# Comparison of the Multivariate Genetic Architecture of Eight Major Psychiatric Disorders Across Sex

**DOI:** 10.1101/2023.05.25.23290545

**Authors:** Ted Schwaba, Travis T. Mallard, Adam X. Maihofer, Mijke Rhemtulla, Phil H. Lee, Jordan W. Smoller, Lea K. Davis, Michel G. Nivard, Andrew D. Grotzinger, Elliot M. Tucker-Drob

**Affiliations:** Department of Psychology, University of Texas at Austin; Population Research Center, University of Texas at Austin; Psychiatric and Neurodevelopmental Genetics Unit, Center for Genomic Medicine, Massachusetts, General Hospital; Research Service, Veterans Affairs San Diego Healthcare System; Department of Psychiatry, University of California, San Diego; Center of Excellence for Stress and Mental Health, Veterans Affairs San Diego Healthcare System; Department of Psychology, University of California, Davis; Stanley Center for Psychiatric Research, The Broad Institute of Harvard and MIT; Department of Psychiatry, Massachusetts General Hospital and Harvard Medical School; Center for Precision Psychiatry, Massachusetts General Hospital; Vanderbilt Genetics Institute, Vanderbilt University; Department of Medicine, Vanderbilt University Medical Center; Department of Psychiatry and Behavioral Sciences, Division of Genetic Medicine, Vanderbilt University, Medical Center; Department of Biological Psychology, Vrije Universiteit Amsterdam; Institute for Behavioral Genetics, University of Colorado Boulder; Department of Psychology and Neuroscience, University of Colorado Boulder

**Keywords:** Psychiatric Disorders, GWAS, Genomic SEM, Sex Differences

## Abstract

Differences in the patterning of genetic sharing and differentiation between groups may arise from differences in biological pathways, social mechanisms, phenotyping and ascertainment. We expand the Genomic Structural Equation Modeling framework to allow for testing Genomic Structural Invariance (GSI): the formal comparison of multivariate genetic architecture across groups of individuals. We apply GSI to systematically compare the autosomal multivariate genetic architecture of eight psychiatric disorders spanning three broad factors (psychotic, neurodevelopmental, and internalizing) between cisgender males and females. We find that the genetic factor structure is largely similar between males and females, permitting meaningful comparisons of associations at the level of broad factors. However, problematic alcohol use loads on psychotic disorders in males but not in females, and both problematic alcohol use and post-traumatic stress disorder load more strongly on internalizing disorders in females than in males. Despite a high between-sex genetic correlation, the neurodevelopmental disorders factor exhibited weaker genetic correlations with psychotic and internalizing factors in females compared to males. Four biobehavioral phenotypes (educational attainment, insomnia, smoking ever, and Townsend Deprivation Index) had significant albeit small sex-differentiated associations with the psychotic factor. As GWAS samples continue to grow and diversify, GSI will become increasingly valuable for studying multivariate genetic architecture across groups.

## Intro

Complex traits often exhibit substantial genetic sharing: many of the variants that are associated with one phenotype are also associated with other phenotypes. In the context of psychiatric disorders, this genetic sharing can be formally modelled in terms of broader multivariate factors (e.g., Psychotic Disorders, Neurodevelopmental Disorders, Internalizing Disorders) that represent transdiagnostic dimensions of genetic risk (Grotzinger et al., 2022; Lee et al., 2019, 2021). To date, however, formal multivariate investigations of genetic sharing have been conducted using data that are aggregated over subgroups (e.g., biological sex, socioeconomic strata, nationality, study; Khramtsova et al., 2023; Mostafavi et al., 2020) that may potentially differ in their genetic architectures, or using data that were obtained from one group to the exclusion of others. Such approaches may mask meaningful differences in the patterning of genetic sharing between groups and limit generalizability of findings.

We introduce Genomic Structural Invariance (GSI), a principled method for comparing multivariate genetic architecture between groups of individuals. GSI tests the similarity of model parameters derived from applying Genomic Structural Equation Modelling (Grotzinger et al., 2019) to summary GWAS data, structuring tests in terms of parsimonious sets that aggregate power between multiple GWAS phenotypes and allow for theoretically meaningful inferences about patterns of group similarity and difference.

We use GSI to compare the multivariate architecture of eight psychiatric disorders across biological sex. Males and females ^1^ differ in presentation, course, prevalence, and comorbidity across a wide spectrum of psychiatric disorders (Abel et al., 2010; Dohrenwend & Dohrenwend, 1976; Merikangas & Almasy, 2020). Recent work has investigated sex differences in associations between common autosomal genetic variants and psychiatric disorders (Bernabeau et al., 2021; Blokland et al., 2022; Duncan et al., 2018; Martin et al., 2021; Silviera et al., 2023; Zhu et al., 2022). This work has found that genetic architecture of individual psychiatric disorders is generally similar across males and females. However, previous work has only been positioned to detect large differences, as stratifying samples by sex reduces power considerably (Khramtsova et al., 2019; 2023). Moreover, the small number of putative sex-differentiated genetic effects among individual disorders have been difficult to interpret or relate to established multivariate genetic factor models. In contrast, GSI estimates sex differences in in the context of a low-dimensional factor model that pools information across multiple cells of a high-dimensional genetic covariance matrix into a smaller set of interpretable parameters, thereby aggregating power across GWAS phenotypes and reducing the burden of multiple testing.

We structure our between-sex comparison of the multivariate genetic architecture of eight psychiatric disorders using a genetic factor model composed of three transdiagnostic factors (Psychotic, Neurodevelopmental, and Internalizing). We first test the comparability of these factors across males and females by examining invariance of each disorder’s loadings on these factors. We then test for sex differences in the genetic variance of these factors, patterns of between-factor genetic associations, and associations between factors and other clinically-relevant biobehavioral phenotypes. Finally, we submit the factors to multivariate sex-by-SNP GWAS to identify genetic variants that have sex-differentiated associations.

## Results

### Overview of Genomic Structural Invariance testing

Genomic Structural Invariance (GSI) is a formal method for comparing multivariate genetic architecture between groups. Many different types of grouping are possible, contingent on the availability of group-stratified GWAS summary statistics, including social exposure (e.g., trauma exposed vs. trauma unexposed), experimental condition (e.g., treated vs. untreated), nation (e.g., Sweden, England, Germany), datasets (e.g., UK Biobank, 23andMe), or forms of ascertainment (e.g., self-report vs. clinician diagnosed). Here, we use sex, among cisgender participants.

GSI involves applying Genomic SEM to a group stratified genetic covariance matrix, S. For *k* GWAS phenotypes measured in *g* groups, a symmetric gk×gk genetic covariance matrix is estimated. In the context of comparing multivariate architecture between sexes, this can be written as the following block matrix:

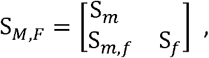

where S_*m*_ and S_f_ are k×k symmetric submatrices containing sex-specific heritabilities on their diagonals and sex-specific genetic covariances among phenotypes off their diagonals, and *S*_*m,f*_ is a k×k asymmetric submatrix containing between-sex within-phenotype genetic covariances on its diagonal and between-sex between-phenotype genetic covariances on the off-diagonal.

GSI uses Genomic SEM (Grotzinger et al., 2019) to model this genetic covariance matrix and compare sets of parameters between groups. Within each group, *g*, a measurement model is specified in which the genetic components of the *k* GWAS phenotypes are specified as linear functions of a smaller set of *m* continuous latent variables as follows:

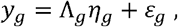

where *y*_*g*_ is a *k* × 1 vector of genetic components of the GWAS phenotypes in group *g, η*_*g*_ is an *m* × 1 vector of latent variables (i.e., factors) in group *g*, Λ_*g*_ is a *k* × *m* matrix of factor loadings relating the latent variables to the genetic components of the GWAS phenotypes in group *g*, and *ε*_*g*_ is a *k* × 1 vector of residuals in group *g*. Also estimated is a *gm* × *gm* latent variable covariance matrix, Ψ, consisting of both within-group covariances and between-group covariances, and a *gk* × *gk* covariance matrix, 0, for the residuals, *ε*.

GSI proceeds according to a sequence of steps in which a researcher tests the equivalence of parameter sets between groups (see **Sequence of Comparison Tests** in the methods section below as well as the Supplementary Note for a plain-language reference guide). First, the equivalence of the Λ_*g*_ parameters (the factor loadings) is tested across groups. To meaningfully compare associations involving a latent variable across groups, it is necessary for (at least some) loadings within Λ_*g*_ to be equivalent across groups. If such (partial) invariance of factor loadings is confirmed, GSI can then be applied to test the between-group equivalence of latent variable variances, equivalence of within-group covariances, and between-group covariances. External variables can be incorporated into the model, such as collateral GWAS phenotypes or SNPs, with tests for group differences in associations between these external variables and each factor.

### Modeling the sex-stratified genetic architecture of psychiatric disorder factors

We curated the most recently available European ancestry sex-stratified GWAS summary statistics for 8 psychiatric disorders **(Table 1):** attention-deficit/hyperactivity disorder (ADHD), problematic alcohol use (ALCH), autism spectrum disorder (AUT), anxiety disorders (ANX), bipolar disorder (BIP), major depressive disorder (MDD), post-traumatic stress disorder (PTSD), and schizophrenia (SCZ). Our selection of GWAS phenotypes followed Grotzinger et al. (2022). However, we did not include compulsive disorders (Tourette’s syndrome, anorexia nervosa, and obsessive-compulsive disorder), as GWAS for these disorders had very low power when sex-stratified. Additionally, we removed AUT in females from the model due to a negative estimate of heritability and potential confounding in GWAS (i.e., high LD score regression (LDSC) intercept inflation relative to GWAS signal).

**Table 1.** Psychiatric Disorder phenotypes used in this study. BIP = Bipolar disorder. SCZ = Schizophrenia. ALCH = Problematic alcohol use. ADHD = Attention Deficit Hyperactivity Disorder. PTSD = Post-Traumatic Stress Disorder. MDD = Major Depressive Disorder. ANX = Anxiety Disorder. Cont. = phenotype measured on a continuous scale. Bin. = phenotype measured on a binary case-control scale. SNP heritability for all phenotypes is scaled in terms of liability.* = For binary GWAS phenotypes, N refers to the sum of effective sample sizes across contributing cohorts, where effective N for each subsample = 4 × (*sample prevalance rate*) × (1 − *sample prevalence rate)* × *Sample Size*. For continuous+ binary traits, N refers to the sum of the continuous sample size and the liability-scale corrected sample size, and can be treated as equivalent to a continuous sample size. See Grotzinger et al., 2023 and Supplementary Note for further details.

| Disorder | Disorder factor | Females |  |  |  | Males |  |  |  | Cont./Bin. | Source/Citation |
| --- | --- | --- | --- | --- | --- | --- | --- | --- | --- | --- | --- |
| | | N* | SNP heritability (SE) | Mean $\chi^2$ | LDSC univariate intercept | N* | SNP heritability (SE) | Mean $\chi^2$ | LDSC univariate intercept | | |
| BIP | PSY | 25,039 | .277(.018) | 1.150 | 0.992(.006) | 19,500 | .314(.025) | 1.137 | 0.997(.007) | Bin. | Blokland et al. (2022) |
| SCZ | PSY | 26,435 | .310(.018) | 1.215 | 0.999(.007) | 37,886 | .386(.016) | 1.339 | 0.986(.007) | Bin. | Blokland et al. (2022) |
| ALCH | PSY, DEV, INT | 68,399 | .027(.007) | 1.024 | 0.990(.006) | 58,337 | .038(.009) | 1.047 | 1.007(.006) | Cont.+Bin. | Walters et al. (2018); Mallard et al. (2022) |
| ADHD | DEV | 13,867 | .138(.028) | 1.121 | 1.060(.008) | 29,628 | .235(.019) | 1.178 | 1.007(.008) | Bin. | Martin et al. (2018) |
| AUT | DEV | 12,806 | -.041(.016) | 1.073 | 1.097(.007) | 35,074 | .183(.016) | 1.127 | 0.987(.007) | Bin. | Grove et al. (2019) |
| PTSD | DEV, INT | 79,846 | .074(.007) | 1.114 | 0.999(.006) | 87,532 | .040(.006) | 1.087 | 1.018(.006) | Cont. | Maihofer et al. (2022) |
| MDD | INT | 145,916 | .051(.004) | 1.130 | 0.992(.007) | 59,315 | .081(.009) | 1.094 | 1.008(.006) | Cont.+Bin. | Blokland et al. (2022); Coleman et al. (2020) |
| ANX | INT | 12,388 | .102(.042) | 1.029 | 1.007(.006) | 7,173 | .206(.063) | 1.020 | 0.991(.006) | Bin. | UKB |

Sex-stratified genetic correlations among disorders **(Figs. 1, S1 & S2)** estimated using Genomic SEM’s implementation of LD score regression (Bulik-Sullivan et al., 2015; Grotzinger et al., 2019) indicated clustered patterns of association that resemble patterns observed in sex-pooled samples (Grotzinger et al., 2022; Lee et al., 2019, 2021). We structured these patterns of genetic covariance in terms of the three correlated factors identified in Grotzinger and colleagues’ (2022) analyses of psychiatric disorders, which used sex pooled data: a psychotic disorders factor (PSY, with indicators SCZ and BIP), a neurodevelopmental disorders factor (DEV, composed of ADHD and AUT, with secondary loadings on MDD and PTSD), and an internalizing disorders factor (INT, composed of MDD, ANX, and PTSD), with ALCH loading on all three factors. This pre-registered (https://osf.io/3rzm6) configural structure fit well (Confirmatory Fit Index= .959; Standardized Root Mean Residual= .080; Fig. 53), and factor loadings from the three factors were highly congruent across sex (Tucker’s Congruence Coefficient= .99, .95, and .94 for PSY, DEV, and INT, respectively; Lorenzo-Seva & Ten Berge, 2006; **Table 52)**, indicating that a three-factor solution was also appropriate for the sex-stratified data. We observed a highly sex-discrepant correlation between ALCH and ADHD (*r*_*g ma1e*_ *=* .24, *r*_*g fema1e*_*=* -.36) and thus allowed for correlated residuals between these two disorders within each sex.

**Figure 1.**
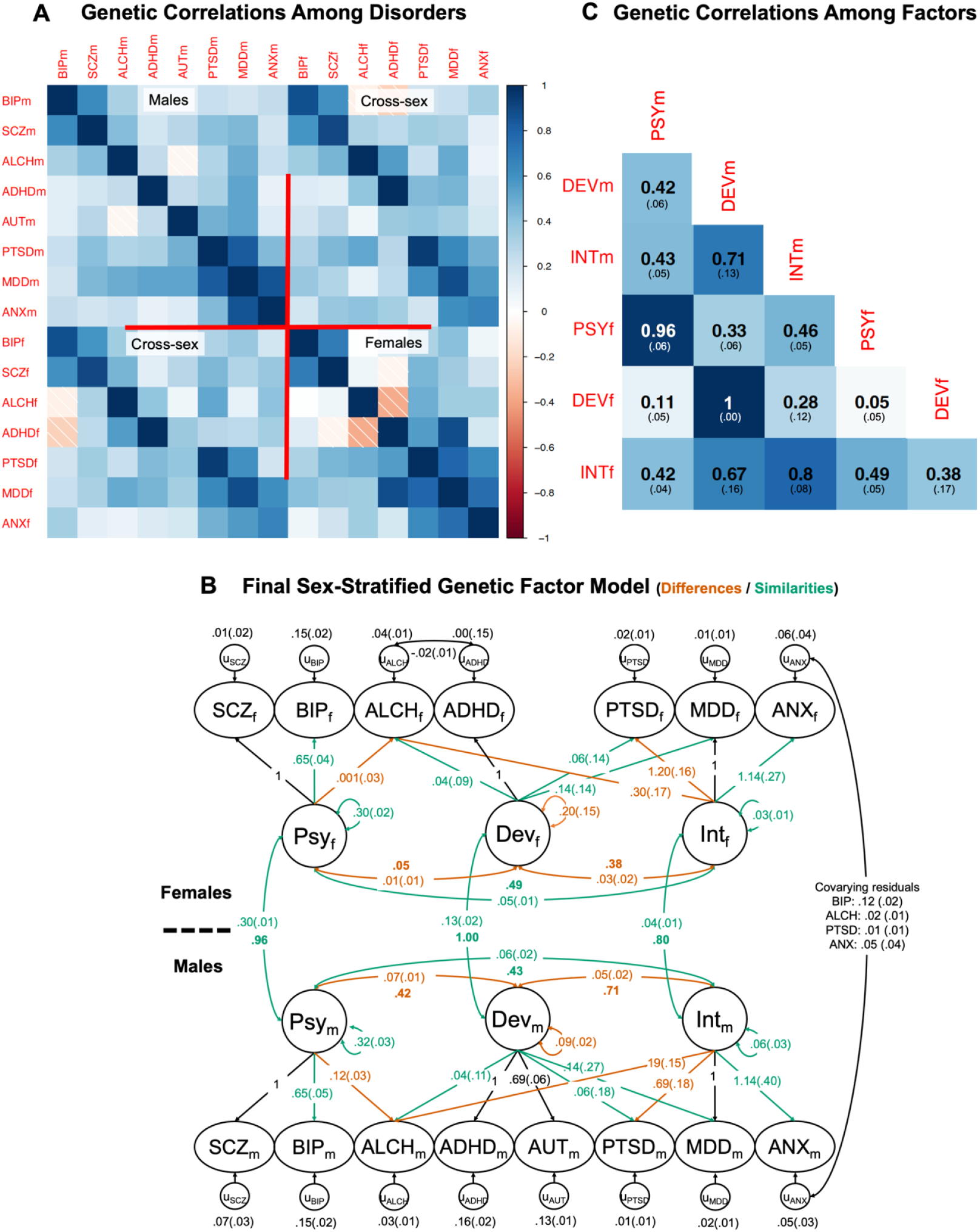
Patterns of sex-stratified genetic associations. Panel A depicts genetic correlations among the eight psychiatric disorders measured in males (subscript m) and females (subscript f). Panel B depicts the final sex-stratified genetic factor model, with constraints imposed according to results of Genomic Structural Invariance tests (see **Table S1** for tabulation of all parameters). In this panel, orange parameters are those that differ significantly and substantially between males and females. Green parameters do not differ between groups, and green factor loadings have therefore been constrained to be equal across groups, enabling meaningful comparison of the higher-order parameters. Unstandardized estimates are represented in plain text outside of parentheses, with SEs inside of parentheses. To facilitate interpretation, standardized covariances (i.e. correlations) among factors are additionally reported as balded parameters. Parameters in black were not tested for sex differences. Loadings of 1, without standard errors reported, represent identification constraints required to set the metric of the factor. Cross-sex cross-factor associations were estimated but not depicted in Panel B; Panel C depicts the full set of genetic correlations among the factors, with SEs inside of parentheses.

### Sex differences in loadings of individual disorders on factors

We implemented a pre-registered two-criterion test of sex differences in model parameters that required parameters to surpass both a Bonferroni-corrected significance threshold and an effect-size threshold for a local measure of parameter difference across groups (Local Standardized Root Mean-square Difference; localSRMD) that we developed specifically for this analysis (see Methods and Supplemental Information for description and validation, and https://rpubs.com/tedooooooooooo/localsrmd for a step-by-step tutorial on how to calculate and apply this index). LocalSRMD is a standardized index of the average group difference within a pre-specified set of parameters of interest, and can be interpreted on a scale similar to a standardized regression coefficient. Relying solely on a statistical significance threshold can result in the detection of trivial differences for high powered comparisons, whereas relying solely on an effect size threshold can result in the detection of differences that are sizable albeit indistinguishable from sampling variation for lower powered comparisons. Complete results of these comparison tests are reported in **Table S3**.

Loadings of SCZ, BIP, and ALCH on the psychotic factor differed between males and females: constraining these loadings to be equal across sex led to a significant decrease in model fit (χ^2^(2) = 9.57, *p* = .008; *p*-value threshold= .0165) that was large in magnitude (Funder & Ozer, 2019) according to local Standardized Root Mean Residual (localSRMD; 0.458, threshold= 0.065). We identified ALCH as the source of this misfit; it was a significant indicator of the psychotic factor in males but not in females (Fig 1). Allowing ALCH to load differentially on this factor in males and females rendered model misfit nonsignificant, thus establishing partial invariance and permitting meaningful between-sex comparisons of associations involving the psychotic factor in subsequent analyses.

Loadings of ADHD, PTSD, ALCH, and MDD on the neurodevelopmental factor did not significantly differ between males and females (χ^2^(3) = 5.85, *p* = .119; localSRMD = 1.21), establishing invariance. These loadings were therefore constrained to be equal across sex for remaining analyses.

Finally, loadings of ALCH, PTSD, MDD, and ANX on the internalizing factor were significantly and strongly differentiated between males and females (χ^2^(3) = 23.28, *p* = 3.5*10^−5^, localSRMD = 2.63). ALCH and PTSD were determined to be the sources of these differences, as relaxing equality constraints across sex for these two factor loadings rendered misfit nonsignificant **(Table S3)**, establishing partial invariance for this factor. In this relaxed model, the unstandardized loadings of both ALCH and PTSD on internalizing disorders were stronger in females than in males. However, for PTSD, this difference did not generalize to the standardized model (.756 in females; .799 in males), which indicates that the difference in unstandardized factor loadings is attributable to higher heritability of PTSD in females than males (LDSC h^2^ = .07 vs.. 04). We carried forward this three-factor, eight-disorder model, with partially constrained psychotic and internalizing factor loadings and fully constrained neurodevelopmental factor loadings (see **Figure 1)**, to all subsequent tests of sex differentiation.

### Testing equivalence of factors across sex

To test whether the genetic architecture of each of the sex-specific factors was equivalent across sex, we fit a simplified model in which the corresponding disorder factors were fully collapsed across males and females (i.e., we modeled three total latent factors, with each indicated by both male-stratified and female-stratified disorders, retaining the partially constrained set of loadings established in the previous step). This model fit significantly worse than the full model that specified sex-specific factors (χ^2^(15) = 59.93; *p* = 2.60*10^−7^), indicating that the genetic factors were not isomorphic across sex. Note that this omnibus test, which draws power from differences in the full set of factor variances and covariances between and within males and females, was considerably more powerful than localized tests that sequentially compare individual factors across sex (cf. van der Sluis et al., 2005). Indeed, the correlations between each of the three corresponding genetic factors measured in males and females were not significantly less than unity (*ps* > .016; **Table S3)**. This may be due to lack of power of the cross-sex genetic correlation to distinguish strong from perfect overlap given currently available sample sizes.

### Sex differences in the genetic variance of factors

We found that the genetic variance of the neurodevelopmental factor was greater in females than in males, as constraining the variance to be equal across sex worsened fit significantly and substantially (χ^2^(1) = 7.97, *p* = .005; localSRMD = 0.317 (threshold= .150); **Table S3)**, reflecting the observation that the genetic factor common to ADHD, PTSD, ALCH, and MDD captures greater genetic variance in females (σ^2^ = .20, se = .15) than in males (whose neurodevelopmental factor also includes AUT; σ^2^ = .09, se = 0.02). Importantly, this sex difference in the genetic variance of the neurodevelopmental factor was driven by the inclusion of AUT (in males) in the model. When AUT was removed from the model, estimates were approximately equal in males and females (in males, σ^2^ = 0.26, se = 0.18, in females, σ^2^ = 0.09, se = 0.02). Genetic variance for the psychotic and internalizing factors did not differ across sex.

### Sex differences in covariance among factors

Covariances of the neurodevelopmental factor with the internalizing and psychotic factors were highly sex-differentiated (χ^2^(2) = 20.07; *p* = 4.4*10^−5^; localSRMD = 2.10 (threshold= 0.100); **Table S3)**. In males, these associations were much stronger (standardized, *r*_*g*_(DEV_m_,PSY_m_) = .42; *r*_*g*_(DEV_m_,1NT_m_)= .71) than in females (*r*_*g*_ (DEV_f_,PSY_f_) = .05, *not significant; r*_*g*_(DEV_f_,INT_f_) = .38; see **Figure 1**. These differences, which were apparent in both the unstandardized and the standardized parameters, indicate a relative differentiation of genetic liability to Neurodevelopmental disorders from the Internalizing and Psychotic disorders in females compared to males. To follow up on this finding, we explored patterns of cross-sex, cross-disorder associations, which provided convergent evidence that the neurodevelopmental factor in females was highly differentiated from the others. For example, the neurodevelopmental factor in females correlated weakly with the psychotic factor in males (*r*_*g*_ = .11) whereas the converse association, between the neurodevelopmental factor in males and the psychotic factor in females, was stronger (*r*_*g*_ = .33; **Figure 1)**.

We investigated the effect of removing AUT completely from the model, rather than estimating it solely in males. Although this afforded 1:1 comparability of neurodevelopmental factor indicators across sex, it paradoxically decreased (from 1 to .86) the between-sex genetic correlation between the male and female neurodevelopmental factors. Moreover, removing AUT in males from the model decreased the genetic correlation between the neurodevelopmental factor in males and all other factors. These findings suggest that the genetic architecture of AUT in males is broadly shared with other disorder factors, within and between males and females. Nevertheless, with AUT removed from the model, the association between the psychotic factor and the neurodevelopmental factor continued to differ by sex (in males, *r* = .20, in females, *r* = .04, χ^2^(1) = 4.00, *p* = .045). We detail these sensitivity analyses in full in Supplemental **Figs. 510 & 511**.

### Sex differences in associations with other biobehavioral phenotypes

Among 24 disorder-relevant biobehavioral phenotypes with sex-differentiated genetic architecture (rgs between males and females ≤.95), four exhibited significantly sex-differentiated associations with the psychotic factor (*p* < 6.6*10^−4^), as illustrated in **Figure 2** and **Table 54:** Insomnia (*r*_*g male=*_ .112, 95% CI [.044, .180]; *r*_*g female*_ = -.048 [-.113, .017]), Townsend deprivation index (*r*_*g male*_ = .288 [.204, .372]; *r*_*g female*_ = .108 [.024, .191]), years of educational attainment (*r*_*g male*_ = .087 [.030, .144]; *r*_*g female*_ = .214 [.044, .180]), and smoking ever (*r*_*g male*_ = .238 [.176, .300]; *r*_*g female*_ = .087 [.022, .152]). Each of these differences remained significant in sensitivity tests that specifically accounted for sex differentiation in the genetic architecture of problematic alcohol use, confirming that this difference reflects factor-level differentiated architecture **(Table 55)**. Compared to past investigations that have identified few sex-differentiated associations involving individual disorders, this result demonstrates the ability of multivariate analyses to recapture power lost by sex-stratification of data. However, none of these differences were substantial in magnitude according to our pre-specified localSRMD cutoff of .150.

**Figure 2:**
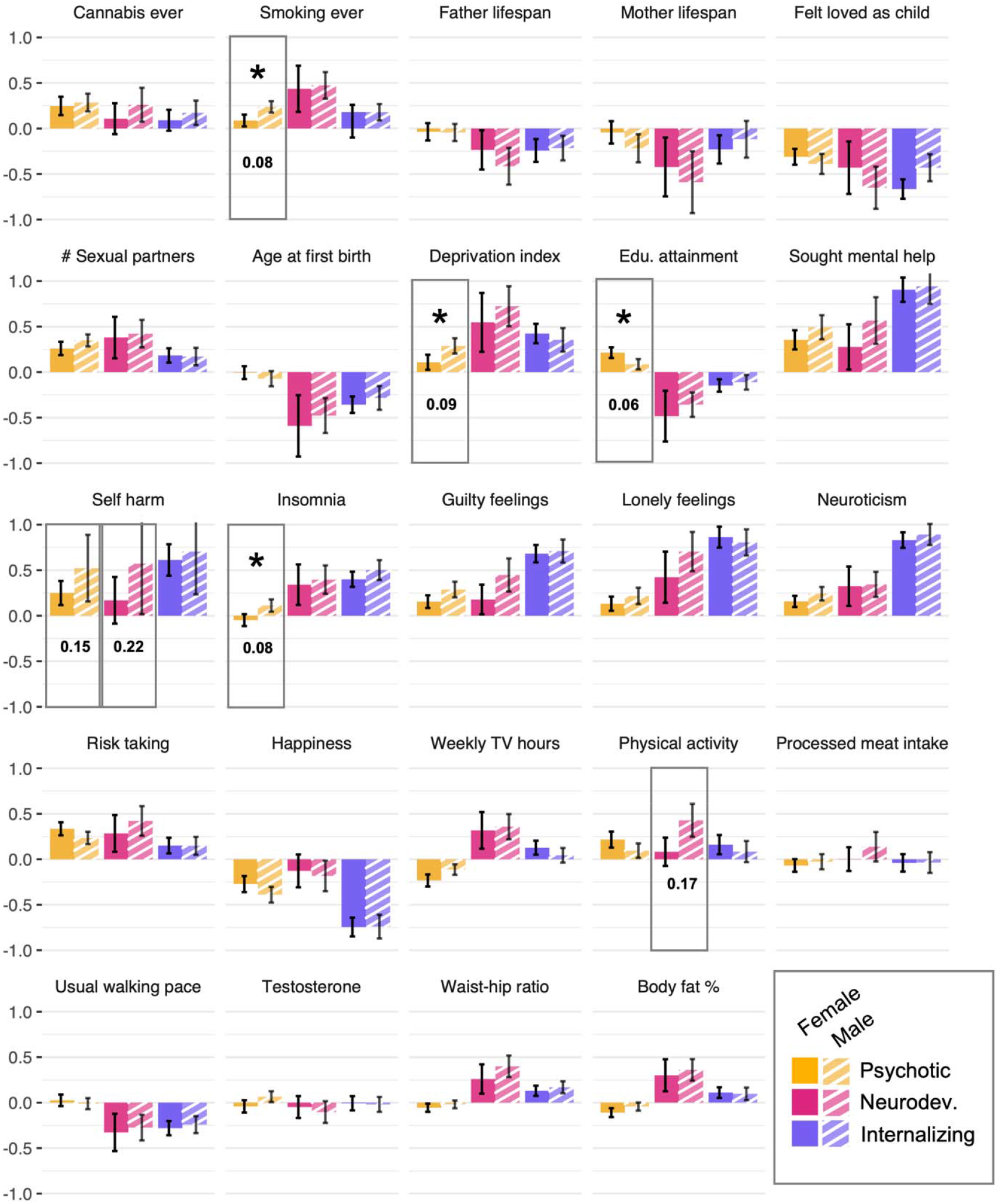
Genetic correlations between disorder factors and relevant biobehavioral phenotypes among males and females. Error bars depict SEs for each association. An asterisk indicates that associations differ between males and females at Bonferroni-corrected *p* < .00066. Numbers below associations indicate localSRMD values for the magnitude of difference and are depicted for associations that with localSRMD ≥.150 or significant differentiation. Boxed associations differ either significantly or with localSRMD ≥.150.

Our analyses indicate that it may also be possible to identify group-differentiated associations between two phenotypes even when one phenotype does not have group-stratified genetic data (e.g., sex-stratified GWAS are available for phenotype A, but only sex-pooled GWAS are available for phenotype B), which would broaden investigations of group differences to a wider variety of phenotypes. Indeed, for educational attainment and smoking, but not insomnia nor deprivation, associations with the psychotic disorder factor were sex-differentiated even when we meta-analytically combined GWAS summary statistics across males and females and examined associations with this sex-pooled GWAS phenotype. In the Supplementary Note, we conduct simulations that identify the conditions under which group-differentiated genetic correlations can be recovered when one of the two phenotypes does not have group-differentiated data.

### Sex x SNP interactions on Multivariate Factors

To test for Sex x SNP interactions at the level of the broad factors, we compared a multivariate GWAS model in which the individual SNP effects on the factor were freely estimated in both males and females, to one in which the effects were constrained to be equal across sex. Of the 4.53 million SNPs available for this analysis, no effects were sex-differentiated at genome-wide significant levels *(ps* ≥ 5*10^−8^) for any of the three factors. However, 15 lead SNPs (5 PSY, 3 DEV, 7 INT) displayed suggestive evidence *(ps* ≤ 5*10^−6^) for sex-differentiated associations **(Tables S8-Sll)**. Full results are reported in the Supplementary Note **(Figures S4-S8)**.

We also submitted each of the sex-stratified disorder factors to multivariate GWAS so as to estimate sex-stratified GWAS of each of the three broad factors. In males, 19 psychotic factor lead SNPs, 0 neurodevelopmental lead SNPs, and 3 internalizing lead SNPs were genome-wide significant. In females, 12 psychotic factor lead SNPs, 0 neurodevelopmental lead SNPs, and 5 internalizing lead SNPs were genome-wide significant **(Figures S4-S8, Tables S15-S16)**. Of this set, two significant lead SNPs for the psychotic factor overlapped across males and females (*r*^2^ > .10), one on chromosome 6 and one on chromosome 12.

Given the general similarity of factor structure across males and females, the high between-sex genetic correlations among the factors, and the lack of genome-wide wide significant sex-differentiated SNP effects on the factors, we believe it currently remains sensible for GWAS discovery to rely on the added power of sex-pooled analyses rather than sex-stratifying discovery samples.

## Discussion

We have described a framework for testing Genomic Structural Invariance, which we applied to compare the autosomal multivariate genetic structure of psychiatric disorder factors across cisgender males and females. Regarding associations between factors and their indicator disorders, we found that psychotic and internalizing factors had different genetic relations with problematic alcohol use by sex. Problematic alcohol use was genetically related to the psychotic disorders factor in males but not females, and was more strongly associated with the internalizing factor in females than in males.

Additionally, in an unstandardized model, PTSD was more strongly related to the internalizing factor in females than in males; comparison with a standardized model revealed that this difference was attributable to greater heritability of PTSD in females. The remaining psychiatric disorders loaded equivalently across males and females on their respective factors, with at least two invariant loadings per factor. This partial invariance of factor loadings indicates that the factor structure of the psychiatric traits examined was generally similar across sex, thus allowing for meaningful comparison of higher-order associations involving the factors.

At the level of between-factor associations, we found that strong patterns of genetic sharing across transdiagnostic factors commonly reported in sex-pooled data may more characteristic of males than females. The positive association between the neurodevelopmental factor and the psychotic factor, which has been identified in past multivariate research on psychiatric disorders (in Grotzinger et al., 2022, *r* = .31), may be driven largely by a positive association among males *(r* = .42), as these two factors were essentially orthogonal among females *(r* = .05). Additionally, the neurodevelopmental factor was more strongly related to the Internalizing factor in males *(r* = .71) compared to females *(r* = .38); in past sex-pooled analyses the correlation between the two was *r* = .54 (Grotzinger et al., 2022). Taken as a whole, these comparison tests indicate that the genetic architecture of the disorder factors is similar, but not identical, across males and females, and that considering factors separately by sex affords the identification of specific fulcrums of sex differentiation.

In contrast with past research, where low statistical power has only enabled identification of large sex-differentiated genetic associations between individual disorders and other biobehavioral phenotypes (Khramtsova et al., 2018), aggregation of power at the factor level allowed us to identify four phenotypes that had significantly sex-differentiated associations with the psychotic disorders factor: educational attainment, insomnia, smoking ever, and Townsend Deprivation Index. However, these differences were small in magnitude (as indexed by localSRMD, standardized differences< 0.150), indicating that, at least for the tested set of phenotypes, shared genetic architecture between biobehavioral phenotypes and disorder factors was not highly discrepant between males and females. Additional evidence for general similarity in the genetic architecture of psychiatric disorder factors across males and females comes from results of sex-by-SNP GWAS analyses, where we identified common genetic variants that had genome-wide significant effects in male-only and female-only samples, but no variants with significantly sex-differentiated effects. Larger sex-stratified GWAS sample sizes will be needed to detect such variants if they exist.

Sex differences in multivariate genetic architecture may emerge via complex interplay of biological and/or social pathways. Biologically, gonadal hormones such as testosterone have been hypothesized to cause sex differences in psychiatric disorders (Gogos et al., 2019), as these hormones operate via relatively distinct pathways in males and females and display strongly sex-differentiated genetic architecture (Sinnott-Armstrong et al., 2021). In this study, however, we did not find that genetic associations between testosterone and disorder factors differed between males and females (Figure 2). Alternately, sex differences in genetic architecture may emerge from social pathways, like living in a gendered society, that differentiate the expression of heritable factors that need not differ biologically between males and females. For instance, the physical changes associated with earlier pubertal development may affect peer relations differently for girls and boys, which may in turn differentially influence mental health across sex (Harden et al., 2014; Moore et al., 2014). Additionally, PTSD is more frequently a result of sexual assault among females and of combat experiences among males (Lake et al., 2023; Tolin & Foa, 2006). If females are more likely to experience sexual assault, and sexual assault (compared to combat exposure) is especially likely to activate genetic risk for internalizing disorders, among other manifestations of psychopathology (Bourgeois et al., 2018; Miller et al., 2004; Miller & Resick, 2007), genetic liability to PTSD may be more heritable in females and load more strongly on an internalizing factor in females than in males, as was observed here. For a final example of how social pathways may influence genetic architecture, ADHD and AUT are underdiagnosed and misdiagnosed at higher rates in females than males (Lai & Baron-Cohen, 2015), and females with autism are more likely to be diagnosed with internalizing disorders than males (Rødgaard et al., 2021), meaning that females with autism may often be included as cases in GWAS of other psychiatric disorders. This may explain, in part, why including AUT as an indicator of the male neurodevelopmental factor increased that factor’s genetic overlap with all other factors in females. Overall, given the wide heterogeneity in potential pathways underlying sex-differentiated genetic architecture, we caution against overconfidence in pinpointing the etiology of the differences that we have identified here.

One important limitation in examining multivariate genetic architecture is that the genetic covariances that serve as the basis for Genomic SEM can be biased by a variety of factors, such as cryptic population stratification and cross-trait assortative mating, although there is some debate about the magnitude of these biases (Border et al., 2022; Grotzinger & Keller, 2022). As we focused on sex differences in genetic covariances, uncontrolled population stratification and assortative mating would need to be asymmetric across sex in order to bias results. A second limitation in this study is that our selection of phenotypes for analysis was constrained by the availability of high-quality GWAS data. This was notable for autism, for which female-stratified data exhibited high levels of genetic confounding, and for anorexia nervosa and Tourette’s syndrome, for which no well-powered sex-stratified GWAS summary data yet exist. As such data become available, future research will be able to expand and refine the inferences drawn here.

While presently applied to the analysis of sex differences, the framework introduced here can readily be applied to compare multivariate genetic architecture for a variety of possible grouping variables, such as social exposure (e.g., trauma exposed vs. trauma unexposed), experimental condition (e.g., treated vs. untreated), nation (e.g., Sweden, England, Germany), datasets (e.g., UK Biobank, 23andMe), or forms of ascertainment (e.g., self-report vs. clinician diagnosed). We have focused here on subgroups sampled from the same general ancestral population. An important extension of GSI for future work will be to allow for comparison of groups sampled from different ancestral populations. In such circumstances, additional provisions will be necessary, such as carefully integrating LD scores from each population with between-population LD scores, so as to estimate within- and between-population components of the genetic covariance matrix (Brown et al., 2016; Turley et al., 2021). Validating such an approach for integration with GSI is an active area of ongoing work.

We have introduced GSI as a formal statistical framework for comparison of multivariate factor structure between groups, and applied it to document important similarities and differences in the genetic architecture of eight major psychiatric traits across sex. Whenever participants are drawn from analytically separable groups, ensuring that the genetic architecture under investigation is comparable across these groups is required to meaningfully interpret their combined analysis. As the range of phenotypes for which large GWAS samples in multiple groups expands, GSI will be a critical tool for formal, principled group comparisons of multivariate genetic structure.

## Methods

### Subgroup-Stratified Genetic Covariances

In the context of subgroup-stratified genetic covariance, *g* separate symmetric k × k genetic covariance matrices (S) can be estimated for *k* GWAS phenotypes measured in *g* groups. Here, we group by biological sex. Thus, we have

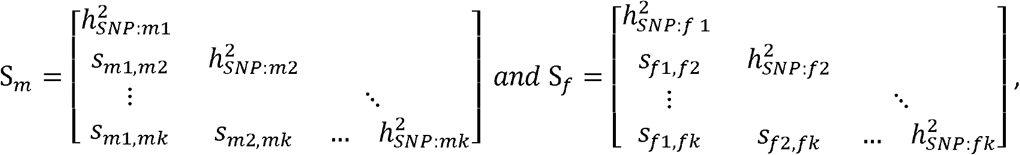

where the diagonal elements 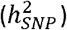 represent the heritabilities of phenotypes 1 through k, the off diagonal elements (*s*_*i,j*_) repesent the genetic covariances (also termed the coheritabilites) among phenotypes *i* and *j*, and the subscripts *m* and *f* represent sex-specific terms for males and females, respectively. Outside of a genomic context, group-specific matrices can be simultaneously modelled using multigroup structural equation modelling. However, in a genomic context, within-trait and between-trait covariances can be computed across groups within the same matrix, i.e.

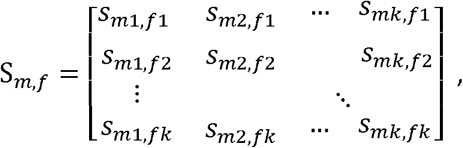

where the diagonal elements contain the between-group within-trait covariances, and off diagonal elements contain the between-group between-trait covariances. (This data structure more closely resembles that observed for paired-sample phenotypic data, such as husband-wife dyads, or longitudinal repeated measures data). Including genetic covariances across multiple groups in a single matrix is possible in Genomic SEM because genetic covariances index similarities in genetic architecture across phenotypes, rather than similarities in phenotypes across individuals. Here, *S*_*m,f*_ is itself k×k, but is not necessarily symmetrical, as the genetic covariance between phenotype *i* in males and phenotype) in females is not necesarily the same as the genetic covariance between phenoytpe *i* in females and phenotype *j* in males. When members of each group are sampled from the same homogeneous continental ancestry (as in the empirical application reported in this paper), estimating the between-group genetic covariances is straightforward: group-specific GWAS phenotypes are simply entered as separate variables when estimating genetic covariances (e.g. with LDSC).

Inclusion of S_*m*_,_*t*_ along with S_*m*_ and S_*f*_ allows for estimation of a single gk×gk genetic covariance matrix, *S*_*MF*_, containing within-group and between-group genetic covariances. Such a matrix can be written in compact form as the following block matrix,

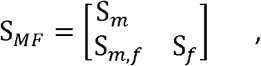

where S_*m*_, S_f_, and S_m_,_f_ are submatrices taking the forms described earlier (see Figure S1 for an example). Below, we provide a principled approach for testing between-group invariance, and lack thereof, of model parameters derived from the application of Genomic SEM to *S*_*MF*_.

### Genomic Structural Invariance (GSI)

Testing sex differences within S_MF_ can be carried out one pair of elements at a time. Such an approach, however, suffers from several limitations including a high multiple testing burden, failure to capitalize on potentially low-dimensional (i.e., factor-level) structure of within-group genetic covariances to enhance power, and lack of direct inferential mapping of results to empirical multivariate models.

Here, we describe a principled multivariate method, Genomic Structural Invariance (GSI), for interrogating sex differences within *S*_*MF*_. Building on classical methods for measurement invariance within structural equation modelling (Horn & McArdle, 1992; Putnick & Bornstein, 2016; Vandenberg & Lance, 2000), GSI tests invariance of model parameters derived from the application of Genomic SEM to *S*_*MF*_, reducing the number of parameters tested to a smaller set that is both theoretically meaningful and aggregates power across multiple cells of *S*_*MF*_.

The genetic covariance matrix, *S*_*MF*_, can be flexibly modelled with Genomic SEM via a user-specified model in which a set of free parameters *(θ)* are estimated by minimizing a fit function that indexes the discrepancy between the model-implied genetic covariance matrix 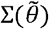 and *S*_*MF*_. Here we estimate use S_MF_ using the multivariable version of LDSC available in the GenomicSEM software, and use the Genomic SEM default of a diagonally-weighted least squares fit function with sandwich corrected standard errors that take into account the sampling covariances among elements of S_MF_, as described in Grotzinger et al. (2019). For *G* groups, a measurement model is specified within each group, *g*, in which the genetic components of the *k* GWAS phenotypes are specified as linear functions of a smaller set of *m* continuous latent variables as follows:

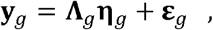

where ***Y***_*g*_ is a *k* × 1 vector of genetic components of the GWAS phenotypes in group g, **η**_***g***_ is an *m* × 1 vector of latent variables (i.e., factors) in group *g*, **Λ**_*g*_ is a *k* × *m* matrix of factor loadings relating the latent variables to the genetic components of the GWAS phenotypes in group *g*, and **ε**_*g*_ is a *k* × 1 vector of residuals in group g. The group-specific vectors **y**_*g*_, **η**_*g*_ and **ε**_*g*_, can each be stacked and the group-specific **Λ**_*g*_ matrix can be concatenated, yielding overall vectors **y, η** and **ε**, and overall matrix **Λ**. The model-implied genetic covariance matrix is then

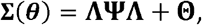

where **Ψ** is an *Gm* × *Gm* latent variable covariance matrix (consisting of both within group covariances and between-group covariances) and **Θ** is a is a *Gk* × *Gk* matrix of covariances among the residuals, **ε**. In **Θ**, residuals of each phenotype are typically allowed to correlate across groups to account for their similarity over and above the latent variables they inform.

We can expand the measurement model to include directed regression coefficients between latent variables with the following addition:

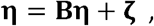

where **B** is a *Gm* × *Gm* matrix of regression coefficients that associate latent variables with each other and **ζ** is a *Gm* × 1 vector of latent variable residuals. This can be particularly useful when including external variables in the model, such as collateral GWAS phenotypes or SNPs.

### Sequence of Comparisons

Genomic Structural Invariance proceeds according to a sequence of comparison tests between groups. To help readers understand this sequence and apply it in their own investigations, we include a brief plain-language reference guide for testing GSI in the supplemental material. Before conducting these tests, the researcher estimates the basic measurement model in Genomic SEM with minimal between-group constraints imposed. After ensuring that the same general configuration of latent factors can be applied across groups, GSI testing can proceed.

As the first step in establishing GSI, the researcher tests the equivalence of the Λ_*g*_ parameters between groups by constraining corresponding parameters to be equal between groups and examining decrease in model fit. This can be done in a fully omnibus fashion, in which all elements of Λ_*g*_ are constrained and tested at once, or for subsets of Λ_*g*_ in turn. Here, we test subsets of Λ_*g*_ corresponding to loadings on each latent factor, which allows us to localize sources of difference to individual factors. In order to meaningfully compare associations involving latent variables between groups, they must be similar in content. That is, at least some estimated loadings within Λ_*g*_ associated with each latent variable must be equivalent across groups (Drasgow, 1984; Meredith, 1993; Putnick & Bornstein, 2016; Vandenberg & Lance, 2000), so that constraining these loadings to be equal does not lead to decreases in model fit beyond a certain prespecified threshold. If the entire set of factor loadings cannot be constrained to be equal across groups, partial invariance across groups may be achieved by systematically freeing the most group-discrepant factor loadings and re-estimating the model until a well-fitting model is identified. If partial invariance cannot be established the latent variable(s) cannot be meaningfully compared between groups -their content differs to an insurmountable extent, and these latent variables can be interpreted independently but not in relation to each other.

After (partial) invariance of factor loadings is confirmed, subsequent tests of GSI can be conducted using a model that retains these between-group patterns of constrained factor loadings. (We note for readers familiar with phenotypic invariance testing that GSI omits testing for invariance of means and intercepts, as such vectors are not defined within genetic space). Here, we conduct four sequential sets of comparison tests that are likely of interest to many researchers examining multivariate genetic architecture: comparing (i) the variances of latent factors between groups, (ii) patterns of covariance among factors between groups, (iii) the correlation between a single factor measured in different groups, and (iv) between-group between-factor patterns of covariance.

External variables may be incorporated into these tests to examine if they demonstrate group-differentiated associations with the latent factors. We illustrate two sets of comparison tests involving external variables: testing whether external GWAS phenotypes have group-differentiated associations with the factors, and testing whether individual common genetic variants have group-differentiated effects on the factors.

### Local Standardized Root Mean squared Difference (localSRMD)

To evaluate the invariance of a set of model parameters across groups, a null hypothesis significance test of their exact equivalence is applied via a nested χ^2^ difference test, in which the χ^2^ for a model in which the parameters of interest are constrained to equality across groups is compared to the χ^2^ for a model in which the parameters within the set are freely estimated within each group. However, a set of parameters that is not exactly equivalent across groups may only differ by a trivial magnitude and thus not indicate a meaningfully large difference. To overcome this, we developed Local Standardized Root Mean Squared Difference (localSRMD), a standardized effect size index of group differences in specific subsets of structural equation model parameters. We have prepared an online tutorial for localSRMD so that researchers can learn more about its calculation and apply it to their own Genomic SEM analyses (https://rpubs.com/tedooooooooooo/localsrmd). LocalSRMD is a modified version of Standardized Root Mean Residual (SRMR) that indexes the average standardized extent to which estimates from a constrained set of parameters of interest within a structural equation model differ from those obtained when the same set of parameters are freely estimated. It is calculated using a similar equation as SRMR (Chen, 2007):

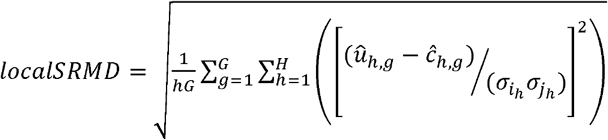

where h is the parameter (variance, covariance, regression, or residual variance) involving (latent or observed) variables *i* and *j* estimated within group *g, H* is the total number of parameters within the set of interest, 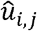 is the estimated value of the parameter in an unconstrained model where the parameters are estimated freely in each group, *ĉ*_*i,j*_ is the estimated value of the parameter in a second model where the parameters are constrained to be equal across groups, and *σ*_*i*_ and *σ*_*j*_ are the estimated standard deviations of *i* and *j* from the unconstrained model, pooled across *G* groups. The *σ*_*i*_ and *σ*_*j*_ terms are estimated as:

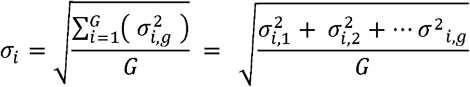

where 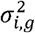 *is* the estimated variance of variable *i* in group *g* and *G* is the number of groups. Note that this approach to calculating localSRMD is unweighted, in the sense that the differences between constrained and unconstrained values in each group contribute equally to its value. This is particularly appropriate for the analysis of sex differences, given the approximately equal proportions of males and females in the general population. Weighted versions of the above equations may be desirable when comparing multivariate genetic architecture between groups whose proportions are unbalanced within the general population.

In effect, localSRMD indexes group differences by calculating the discrepancy between the unconstrained and constrained parameter estimates, standardizing this quantity by dividing by the group-pooled parameter standard deviations, squaring this quantity to remove sign, and averaging across the number of parameters in the set. LocalSRMD is calculated without respect to sample overlap and item intercepts, so it can be easily applied to genomic models. Unlike global fit measures such as Comparative Fit Index (CFI) and SRMR, localSRMD is not calculated using information from the entire model, allowing researchers to test invariance in a localized subset of model parameters instead. As we illustrate through simulation in the Supplementary Note **(Figure S13)**, if a small set of parameters differs substantially between groups within a larger model that is otherwise invariant across groups, the portion of the model that is well-fitting serves as “ballast,” downwardly biasing change in CFI and SRMR when conducting measurement invariance tests (see also Shi et al., 2018). As such, a researcher using those global fit indices may not detect the substantial misfit in part of the model, whereas localSRMD is not affected by these ballast parameters. Also, because localSRMD is a standardized index of misfit, it is similar in scale to a standardized regression coefficient. Researchers can therefore decide cutoffs for localSRMD misfit in the context of their own research questions, rather than relying on suggestions for misfit cutoffs developed in the context of specific simulation examples (Chen, 2007; Cheung & Rensvold, 2002) that may not generalize. For example, in the case of associations between disorder factors and external phenotypes, we pre-registered that localSRMD differences ≥.150 would constitute a meaningfully large effect (e.g., if the internalizing factor correlated .30 with phenotype A in females, and .60 with phenotype A in males, the sex-constrained correlation would be around .45, yielding a localSRMD of 0.15). As described in the Genomic Structural Invariance Testing section below, our approach to measurement invariance employs a two-criterion test in which we require parameters to be both significantly different (as indicated by null hypothesis significance testing) and meaningfully different (exceeding a prespecified localSRMD threshold).

### Sample

We curated the largest and most recent sex-stratified GWAS summary data from individuals of European ancestry for eight major psychiatric disorders **(Table 1)**. We refer readers to the citations in Table 1 for complete details on each ascertainment and quality control procedures for each set of summary data. In these samples, chromosomal sex was compared against self-reports, and participants were included when self-reports of male or female matched their chromosomal sex pattern of XV or XX. In the Supplementary Note, we provide further information on each phenotype, including meta-analytic aggregation of MDD and ALCH data across sources and sex-stratified analyses for ALCH and PTSD that were conducted specifically for this study.

### Factor Model

Genomic SEM was applied to model the sex-stratified genetic covariance structure of 8 major psychiatric disorders (Grotzinger et al., 2019). The liability-scale genetic covariance matrix and its associated sampling covariance matrix was estimated using the *ldsc* function within GenomicSEM software, with LD scores estimated using the European ancestry samples of the 1000 Genomes Project 3v5. For binary phenotypes, we used the sum of effective sample size for N and set the sample prevalence equal to .50, as per new best practices described by Grotzinger et al. (2023). For population prevalence, we used the sex-stratified US population prevalence rates for each psychiatric disorder reported in Martin et al. (2021; **Table 517)**.

We estimated a pre-registered genomic structural equation model that describes associations among these phenotypes in terms of six disorder factors (three per sex): a psychotic factor, neurodevelopmental factor, and internalizing factor, following the structure identified in Grotzinger and colleagues’ (2022) analysis of these phenotypes. Code for all models is available at https://osf.io/wya8p Estimation of this model required smoothing of the covariance matrix to the nearest positive definite matrix; comparisons of pre- and post-smoothed correlations among variables suggests that this did not substantially affect the model **(Figure 514)**. We made two post-hoc modifications to this model: adding a correlated residual between ADHD and ALCH in females and removing AUT in females from the model. These changes and their effects are described in full in the Supplementary Note.

To identify each disorder factor while still allowing for differential factor variance, we constrained each factor’s canonical indicator (SCZ for the psychotic factor, ADHD for the neurodevelopmental factor and MDD for the internalizing factor) to 1. We selected these canonical disorders based on a combination of theory and findings from Grotzinger et al. (2022), and these decisions were supported by similar patterns of covariance for canonical phenotypes across sex. The path diagram of this basic model with no constraints is presented in **Figure S3**, and parameter estimates are included in **Table S2**. The overall fit of this basic, unconstrained model allows us to evaluate the extent to which the number of factors and patterns of loadings on factors are commensurate across groups.

### Genomic Structural Invariance Testing

To test whether the patterns of genetic covariance among the disorders that constitute each factor were similar across sex, we compared the fit of the basic measurement model, described above, with the fit of a constrained measurement model in which a factor’s loadings were constrained to be equal across sex. This constrained model was nested within the basic model, permitting us to directly compare their fits to the data (Vandenberg & Lance, 2000). We assessed change in model fit using two criteria: statistical significance (p-value of a χ ^2^ nested model comparison test, with Bonferroni-corrected threshold of *p* < .016) and magnitude of change (localSRMD, with pre-registered threshold determined through simulation of ≥.060; see pre-registration for simulation results). For each set of parameters, both tests must surpass the prespecified critical values in order for us to conclude sex-differentiated genetic architecture. This strategy allowed us to account for differential sample size across factors: for tests with larger samples, estimates of change in fit were especially precise, so it was important to set a smallest effect size of interest for significant decrements in fit (see Lakens et al., 2018).

After establishing the comparability of factor loadings across groups, we next tested for sex differences in the genetic variance of each factor. To do this, we again conducted nested model comparison tests, comparing the fit of a model with factor variances estimated separately in males and females with the fit of a constrained model in which a given factor’s variance is constrained to be equal across sex. The *p*-value threshold for change in model fit was set at .016 (.05 / 3 factors) and the localSRMD threshold was set at .150.

Third, we tested for sex differences in the within-sex genetic covariances among the disorder factors. We compared the fit of the basic model, where genetic covariances among disorders were estimated freely in males and females, with a constrained model where all covariances with a factor (i.e. for the psychotic factor, cov(PSY, DEV) and cov(PSY, INT)) were constrained to be equal across males and females. The *p*-value threshold for change in model fit was set at .016 (.05 / 3 factors) and the localSRMD threshold was set at .100. We also explored between-factor between-sex genetic patterning, though we did not conduct hypothesis tests of these differences.

Fourth, we tested between-sex genetic correlations for each disorder factor. To do this, we compared the fit of two models: the basic measurement model, where each factor’s genetic correlation between males and females was freely estimated, and a constrained model in which a factor’s genetic correlation between males and females was set to 1. We compared the fits of these models using a 1 *df* χ ^2^ test with a Bonferroni-corrected *p*-value threshold of .016.

Fifth, we examined sex differences in the genetic correlations between factors and 24 external biobehavioral phenotypes (See **Figure 2** and **Table S4)**. To do this, we estimated extensions of the SMF matrix and basic measurement model that included additional sex-stratified phenotype variables. We compared the fit of a freed model, where the genetic covariances between the external phenotype and all three factors were estimated separately among males and females, with a constrained model, where the genetic covariance between a factor and the external phenotype was constrained to be equal for males and females. For these comparisons, we set the Bonferroni-corrected *p*-value threshold at .00066 and the localSRMD threshold at .150. For the external phenotypes where we identified significant sex differences in factor-outcome associations, we meta-analytically collapsed data across sex and estimated additional models on this sex-pooled data in order to test whether sex differences in associations could be identified even with group-aggregated external phenotype data.

### Multivariate GWAS of sex x SNP interaction effects

We conducted specific tests of sex-differentiated genetic architecture at the Single Nucleotide Polymorphism (SNP) level, including the set of 4.53 million SNPs catalogued in the 1000 Genomes Project 3v5 that were present across the 15 sets of phenotype summary statistics. This entailed estimation of two extensions of the basic measurement model that included individual SNPs: one where each latent disorder factor was regressed on the SNP simultaneously, with associations estimated freely, and one where each latent factor was regressed on the SNP simultaneously, with regression parameters of a single factor on the SNP constrained to be equal in magnitude across males and females. For each SNP and each factor, we compared the fits of these two nested models using a 1 *df* χ ^2^ test with a genome-wide significance threshold (5 × 10^−8^).

We then submitted summary statistics obtained through these analyses to FUMA (Watanabe et al., 2017) and estimation of univariate and bivariate LDSC in Genomic SEM. We identified lead SNPs at a relaxed significance threshold (5 × 10^−6^) that were independent at *r* ^2^ < .10 within 250kb. For each of the three multivariate factors, GWAS summary statistics for sex-stratified factors and tests of sex differentiation are available for download at https://osf.io/spg7f/

## Data Availability

GWAS summary statistics from this study are available for download at https://osf.io/spg7f/
Sex-stratified summary statistics used in this study come from the following sources (see Supplementary Note for complete details):
Schizophrenia, Bipolar Disorder, Major Depressive Disorder(1): https://doi.org/10.1016/j.biopsych.2021.02.972
Problematic Alcohol Use(1), ADHD: https://doi.org/10.1016/j.biopsych.2020.12.024
Autism: https://doi.org/10.1038/s41588-019-0344-8
Anxiety and Major Depressive Disorder(2): http://www.nealelab.is/uk-biobank
PTSD: https://doi.org/10.1016/j.biopsych.2021.09.020
Problematic Alcohol Use(2): https://doi.org/10.1176/appi.ajp.2020.20091390

## Footnotes

1 In this paper, we use the term males to indicate people with XY sex chromosomes who identify as male and females to indicate people with XX chromosomes who identify as female. As relatively few nonbinary and transgender people and people with other patterns of sex chromosomes (e.g., **XXY)** are included in genomic studies, we were unable to incorporate these groups in the present research. This necessarily limits the generalizability of our conclusions. However future genetic research on sex and gender will benefit from thoughtfully and meaningfully including people with a diverse set of gender identities and biological chromosomal patterns.

