## Supplementary Note for "Comparison of the Multivariate Genetic Architecture of Eight Major Psychiatric Disorders Across Sex"

**Deviations from pre-registration**

There were six deviations from pre-registered analyses. Regarding phenotype inclusion, we (1) removed AUT in females from the model due to lack of genetic signal (we provide further information in the main text and sensitivity analyses below) and (2) estimated PTSD solely using summary statistics from analyses of continuous data, as opposed to meta-analyzing continuous and case-control data together as pre-registered (see Further Information on Phenotypes below for complete details). Additionally, (3) we excluded one external biobehavioral phenotype from analyses, cardiac problems, as the LSDC-estimated heritability of this phenotype was negative. Regarding modeling procedure, (4) we identified latent variables by setting a single anchor loading per factor to one, as opposed to setting factor variance in females equal to one as pre-registered. This change, which was necessary for model convergence, alters the scaling of parameters but does not substantively change model estimates. (5) We became aware of work within structural equation modeling indicating that an omnibus test of factor equivalence is more appropriate than a test of the distinction of their correlation from *r* = 1.0 (van der Sluis et al., 2009). As a result, we included an omnibus test of the equivalence of factors (see test 2a in the reference guide below and Testing Equivalence of Factors Across Sex in the results section of the main text) and reordered our pre-registered hypothesis tests to include this addition. Finally, (6) we added a correlated residual parameter to our models to account for the negative correlation between ADHD and ALCH in females (see below for sensitivity tests).

**Further Information on GWAS Phenotypes**

As described in the pre-registration for this study, we curated sex-stratified summary statistics for eight of the 11 disorders studied in Grotzinger and colleagues’ (2022) investigation of the multivariate genetic architecture of psychiatric disorders. We excluded three disorders in Grotzinger and colleagues (2022) from this study, anorexia nervosa, obsessive-compulsive disorder, and Tourette’s syndrome, due to a lack of suitable sex-stratified GWAS summary statistics. Results of the univariate LSDC regression for each of the eight phenotypes can be found in **Table 1** in the main text.

The majority of data used in this study came from published studies. SCZ, BIP, and MDD data came from Blokland and colleagues (2022). ALCH data came from Walters and colleagues (2018), provided to us by the authors of Martin and colleagues (2021). We estimated sample size for this phenotype from minor allele frequency using the method in Grotzinger and colleagues (2023). ADHD data came from Martin and colleagues (2018), provided to us by the authors of Martin and colleagues (2021). AUT data came from Grove and colleagues (2019), with sample size estimated using minor allele frequency. ANX and additional MDD data came from the sex-stratified European ancestry analyses of self-reported anxiety (#20002_1287) and depression (#20002_1286) in the UK Biobank, which are publicly available from the nealelab website (<http://www.nealelab.is/uk-biobank>).

Additional ALCH data were contributed to this study by T.T.M., who conducted sex-stratified analyses of problematic alcohol use in the UKB sample, as originally studied in Mallard and colleagues (2022). Specifically, T.T.M. first computed a standardized composite score from items 4-10 of the Alcohol Use Disorder Identification Test, which measure alcohol use problems. Items 1-3 were excluded, as these items solely measure alcohol consumption and are less strongly genetically correlated out-of-sample with alcohol use disorder (Mallard et al., 2022). T.T.M. then used PLINK v2.00 (Chang et al., 2015) to conduct genome-wide analyses on this phenotype, in the same manner as described in the above publication.

PTSD data were contributed by A.X.M., who conducted sex-stratified analyses of PTSD separately by assessment method (continuously-scored symptom counts and case/control diagnosis), as originally studied in Maihofer and colleagues (2022). Specifically, A.X.M. conducted genome-wide analyses on both the continuous PTSD phenotype (N_males_= 87,532, N_females_= 79,846) and case-control PTSD phenotype (N_males_= 2,516 cases/7,975 controls; N_females_= 1,493 cases/4,113 controls) in the same manner as described in the above publication. In preparation for meta-analyzing these results together, we found strong negative genetic correlations between case-control PTSD and other phenotypes, including continuous PTSD (*r*_g_ < -1.00). Given that the vast majority of the PTSD sample contributed continuously-measured data, we dropped the case-control summary statistics from all subsequent analyses.

For both ALCH and MDD, we meta-analytically combined data across two sources (one binary GWAS trait and one continuous GWAS trait) to increase sample size following the method for combined GWAS meta-analysis of continuous and binary traits described in Grotzinger et al. (2023) using a sample size weighted approach wherein the binary trait is weighted by the liability-scale corrected sample size and the continuous trait weighted by the continuous sample size. This approach relies on the assumption that the continuous and binary GWAS traits both represent different approaches to measuring the same continuous trait, such that liability-scale heritability of the binary variable is on a comparable scale to the heritability of the continuous variable. The combined sample size is calculated as the sum of the continuous sample size and the liability-scale corrected sample size, i.e.: $\left( \sum n_{kcont} \right)+\frac{\phi^{2}}{{4P}^{2}\left( 1-P \right)^{2}}\left( \sum{EffN}_{kbin} \right)$ and the combined meta-analysis is treated as continuous when entered into LDSC (see Grotzinger et al., 2023 for explanation of terms in this formula).

**Reference Guide for Testing Genomic Structural Invariance**

Below is a guide to help you apply GSI beyond this study. We also provide an online tutorial for computing and understanding the localSRMD index of model fit, which also demonstrates tests 1 and 1a below, at <https://rpubs.com/tedooooooooooo/localsrmd>.

| **Test** | **Pre-Requisite** | **Parameters Constrained** | **Fit Indices** | **Logic** |
| --- | --- | --- | --- | --- |
| 0) Overall model configuration | N/A | None | CFI & SRMR | Before conducting GSI tests, ensure that the measurement model converges and fits acceptably to the data according to CFI and SRMR. If model fit is unacceptable before any parameters are constrained across groups, the researcher may opt to fit separate measurement models in each group and forego group comparison testing. |
| 1) Full equivalence of factor loadings | N/A | All factor loadings | Nested χ² test (significance) & localSRMD (effect size) | Test whether the patterning of associations among the factor’s indicators (i.e., factor loadings) is equivalent across groups using a χ² test and localSRMD. If the χ² test is nonsignificant OR localSRMD is below the effect size cut-off, the content of the latent factor is meaningfully comparable across groups, and proceed to tests 2-6, retaining constrained loadings across groups. If there is significant AND substantial misfit, the patterning varies between groups; proceed to test 1a. |
| 1a) Partial equivalence of factor loadings | Fail Test 1 | Some factor loadings | Nested χ² test & localSRMD | Equate *some* of the factor loadings across groups while freely estimating others and examine fit indices. If equating even a single estimated factor loading across groups leads to significant AND substantial misfit, the factor’s content and interpretation differs between groups, and cross-group comparisons involving the factor’s variance and covariances are not meaningful. If some loadings can be equated across groups without leading to significant and substantial misfit, constrain those loadings across groups when proceeding. |
| 2a) Omnibus equivalence of factors | Pass Test 1 or 1a | None (compare with simplified model) | Nested χ² test | Estimate a simplified factor model in which corresponding latent factors are collapsed across groups (i.e. for a 1 factor model, estimate one factor for G groups instead of G factors for G groups). Loadings established as invariant in 1/1a should continue to be fixed across groups. If collapsing the factor(s) across groups leads to significant misfit compared to 1/1a, the overall genetic architecture of the factor differs between groups. Specific sources of difference are probed further below. |
| 2b) Associations of corresponding factors across groups | Pass Test 1 or 1a | Factor covariances | Nested χ² test | Returning to the group-stratified factor model established in tests 1/1a, set the covariance between each corresponding factor in different groups equal to its maximum possible value (e.g., *r* = 1.0). If doing so leads to significant misfit, that factor is correlated less than perfectly between groups, meaning the overall genetic architecture of the factor differs between groups. |
| 3) Equivalence of factor variances | Pass Test 1 or 1a | Factor variances | Nested χ² test & localSRMD | Equate the variance of a factor across groups. If doing so leads to significant AND substantial misfit, the variance of that factor differs between groups, meaning the shared genetic architecture among the factor’s indicators accounts for a differential amount of genetic variance across groups. |
| 4) Equivalence of within-group cross-factor covariances | Pass Test 1 or 1a & model multiple factors per group | Factor covariances | Nested χ² test & localSRMD | Equate associations (correlations or covariances) between factors within each group. If doing so leads to significant AND substantial misfit, the association between those factors differs between groups, meaning the degree of shared genetic architecture among those factors differs between groups. |
| 5) Equivalence of cross-group cross-factor covariances | Pass Test 1 or 1a & model multiple factors per group | Factor covariances | Nested χ² test & localSRMD | Equate associations between pairs of factors across groups. If, for example, constraining *cov*(Factor1_group1_, Factor2_group2_) equal to *cov*(Factor2_group1_, Factor1_group2_) leads to significant AND substantial misfit, the association between those two factors differs between groups, meaning the degree of shared genetic architecture across those two factors differs between groups. |
| 6) Equivalence of covariances with external variables | Pass Test 1 or 1a & external variables added to model | Factor covariances | Nested χ² test & localSRMD | Estimate an expanded model that includes additional variables (such as a SNP or a phenotype). Set the association between a variable and a factor in one group equal to the association between that variable and that factor in other groups. If doing so leads to significant and/or substantial misfit, the association of that factor with the external variable differs between groups, meaning that outcome and that factor share differential genetic architecture across groups. |

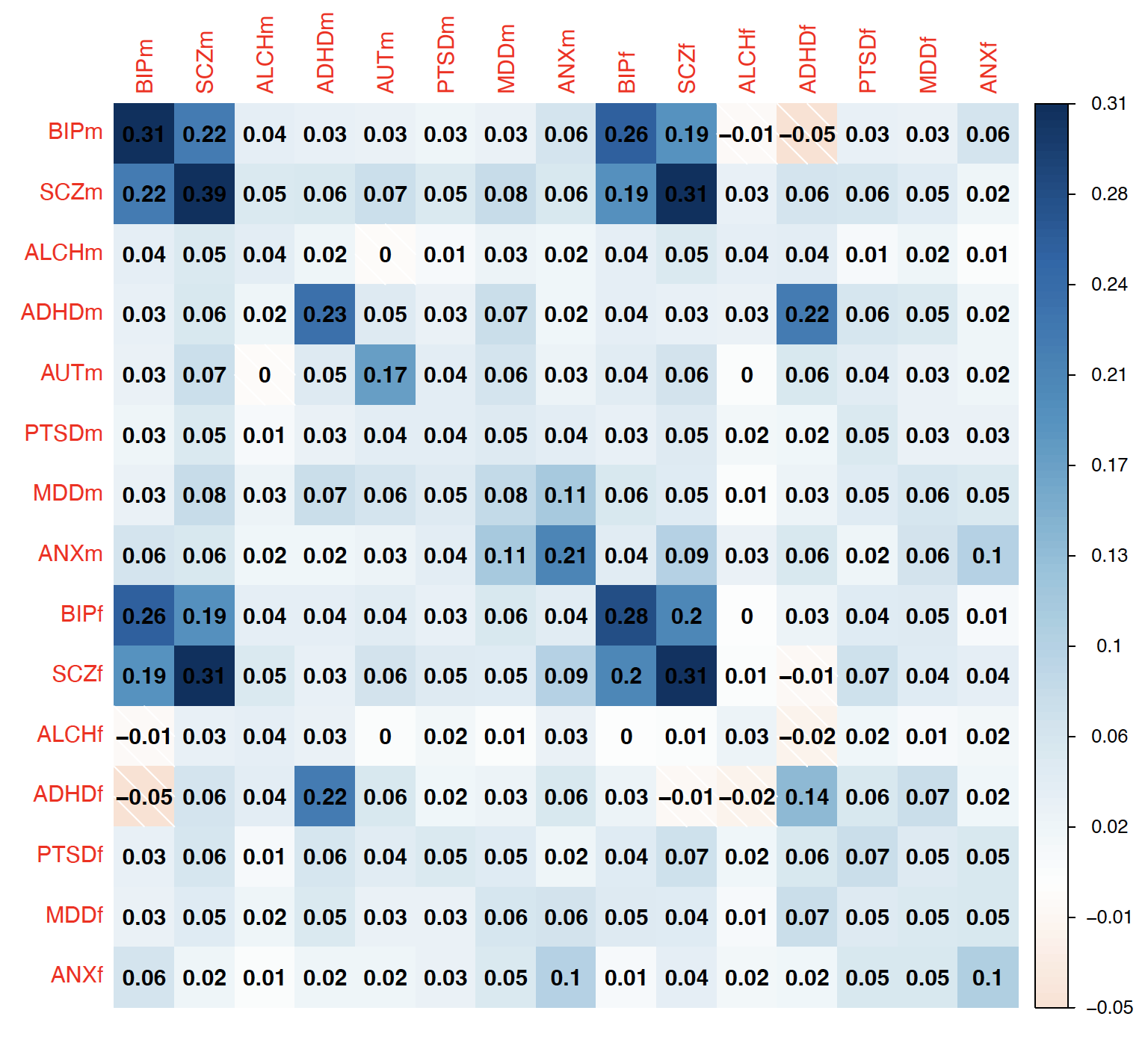

**Figure S1. LDSC Genetic covariance matrix.** Heritability of each phenotype is presented on the diagonal. Disorders with a subscript m indicate males, and disorders with a subscript f indicate females.

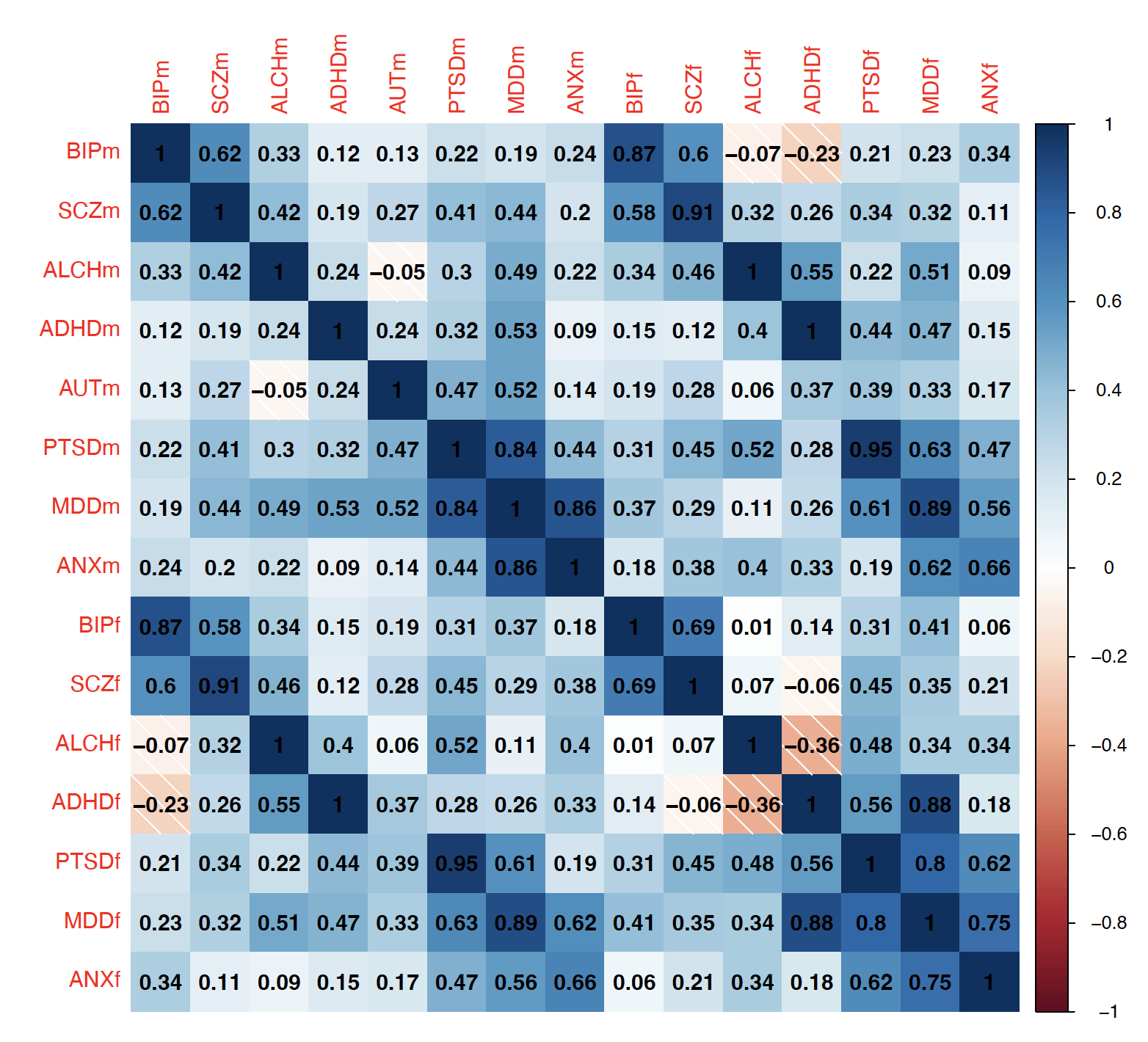

**Figure S2. LDSC Genetic correlation matrix.** Disorders with a subscript m indicate males, and disorders with a subscript f indicate females.

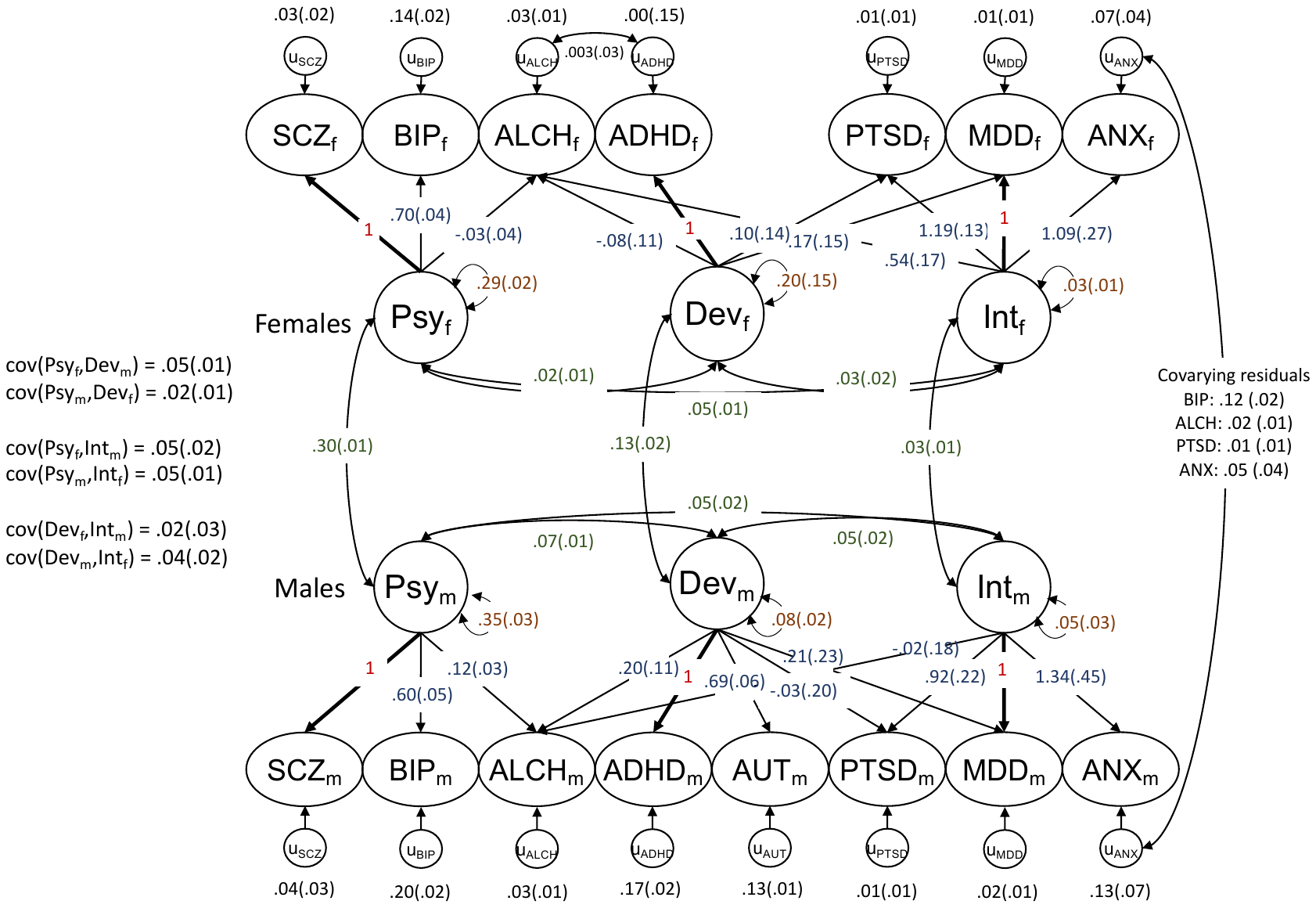

**Figure S3. Measurement model with factor loadings unconstrained.** Red factor loadings are constrained to 1 to identify the model. Blue parameters represent factor loadings, scarlet parameters represent factor variances, and green parameters represent factor covariances. Cross-sex cross-factor covariance parameter estimates are depicted on the left but omitted from the visualization for simplicity. Full parameter estimates are in **Table S2** of the supplementary spreadsheet, and results from the final constrained model are in **Figure 1**. All estimates displayed are unstandardized.

**Sex x SNP interaction effects**

We compared multivariate GWAS models in which each of 4.53 million SNPs predicted factors independently versus equally across sex. These analyses did not reveal any SNP effects that were sex-differentiated at a genome-wide significance threshold (*p*s ≥ $5\times{10}^{-8}$), although for 15 lead SNPs (5 psychotic, 3 neurodevelopmental, 7 internalizing) there was evidence for differentiation at a suggestive significance threshold (*p*s < $5\times{10}^{-6}$; **Figure S4-S6; Table S8-S14**).

For 11 of these 15 suggestive lead SNPs, sex-differentiation was the result of effects in discordant directions across males and females. For the remaining four suggestive lead SNPs, effects were significant at *p* < $5\times{10}^{-6}$in males and near-null in females. Across SNPs, the mean χ^2^ corresponding to sex-by-SNP interactions in associations with each broad factor was greatest for the internalizing factor (1.041), followed by the psychotic (1.020) and neurodevelopmental (0.945) factors, which is consistent with the pattern observed for cross-sex genetic correlations.

Of these 15 suggestive lead SNPs, 5 involved the psychotic factor. Of these five, one association was novel. Three replicated suggestive findings from a previous MTAG analysis of sex-differences in SCZ, BIP, and MDD (Blokland et al., 2022; **Figure S7**), illustrating convergence between participant-level interaction analyses in that study and genomic SEM analyses in this study, which used similar multivariate phenotypes. The fifth suggestive lead SNP, rs77325285, is a locus near SLC45A4 that has been associated with BIP in sex-pooled analyses (Psychiatric GWAS Consortium Bipolar Disorder Working Group, 2011); our findings indicate that risk conferred by this SNP may be sex-differentiated.

Furthermore, of these 15 suggestive SNPs, 3 involved the neurodevelopmental factor (each novel) and seven involved the internalizing factor (six of them novel). The seventh internalizing SNP, rs4360109, has previously been linked to depression and bipolar disorder in single-disorder sex-pooled analyses (Howard et al., 2019; Coleman et al., 2019; Hyde et al., 2016). Our findings indicate that this SNP may confer risk solely in males (*p* = 3.01 $\times{10}^{-7}$), as its association with the internalizing factor in females was near-null (*p* = .649). Across all three factors, no genome-wide significant hits from a sex-pooled GWAS reported by Grotzinger and colleagues (2022) displayed any evidence for sex-differentiation (**Figure S8**).

**LDSC of sex-differentiated effects**

We also submitted these sex-differentiated results to univariate and bivariate LDSC. We identified significant LDSC slopes for sex differences in the psychotic factor (*p* = .010) and the internalizing factor (*p* < .001), but not the neurodevelopmental factor, providing evidence for polygenic signal of sex-differences in these two factors across common genetic variants. We note that although the LDSC slope can be estimated, SNP heritability is not defined for this analysis. No LDSC intercepts were significantly different from 1.0, indicating little residual confounding due to population stratification (PSY = .999, DEV = .964, INT = 1.011). Finally, sex differences did not display significant genetic covariance across factors (*p*s .051 - .160), indicating that sex differentiation occurred in different loci for different factors.

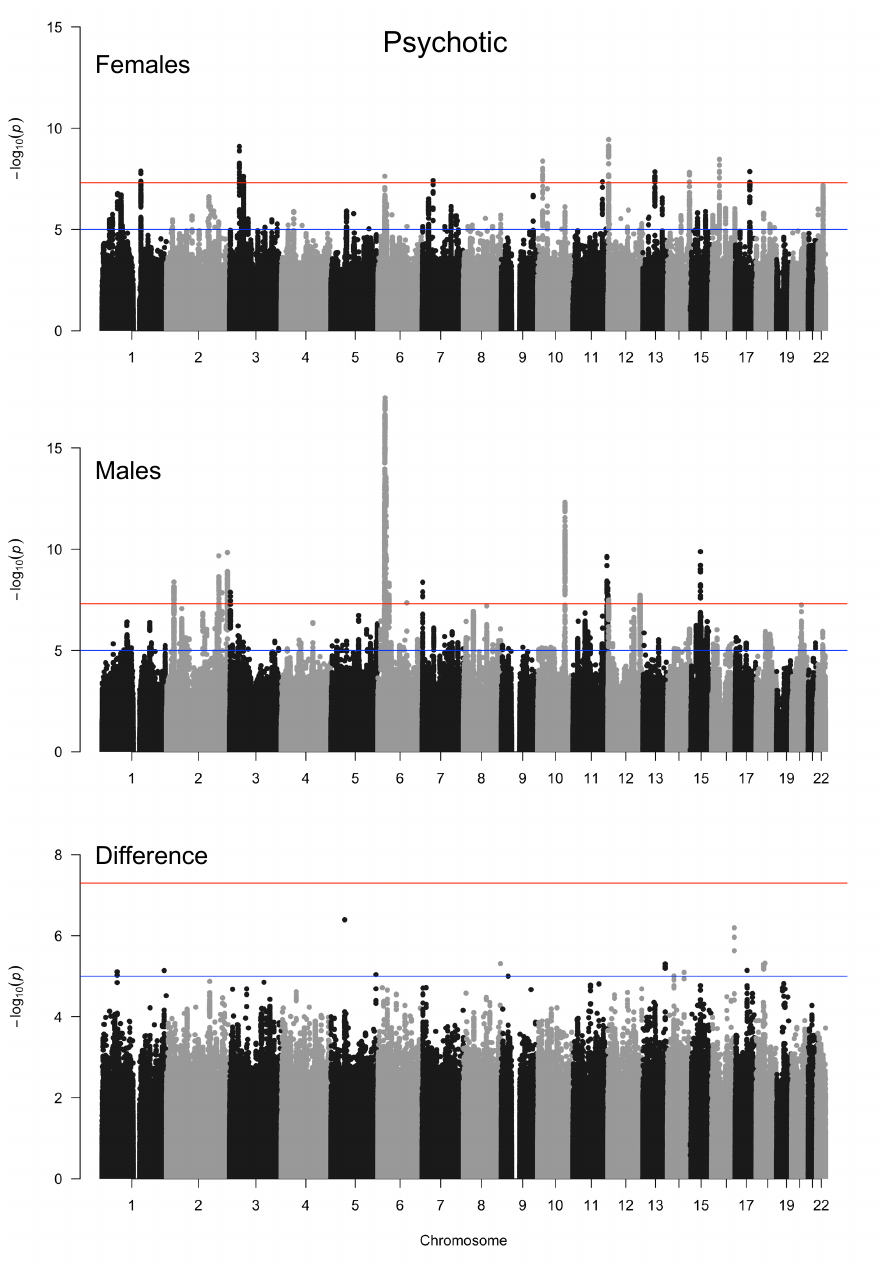

**Figure S4. Manhattan plots of sex differentiation in Psychotic factor associations across SNPs.** The red line denotes the threshold for genome-wide significance ($5\times{10}^{-8})$ and the blue line denotes the threshold for suggestive genome-wide associations ($5\times{10}^{-6}$). Data were available for 4.53 million SNPs.

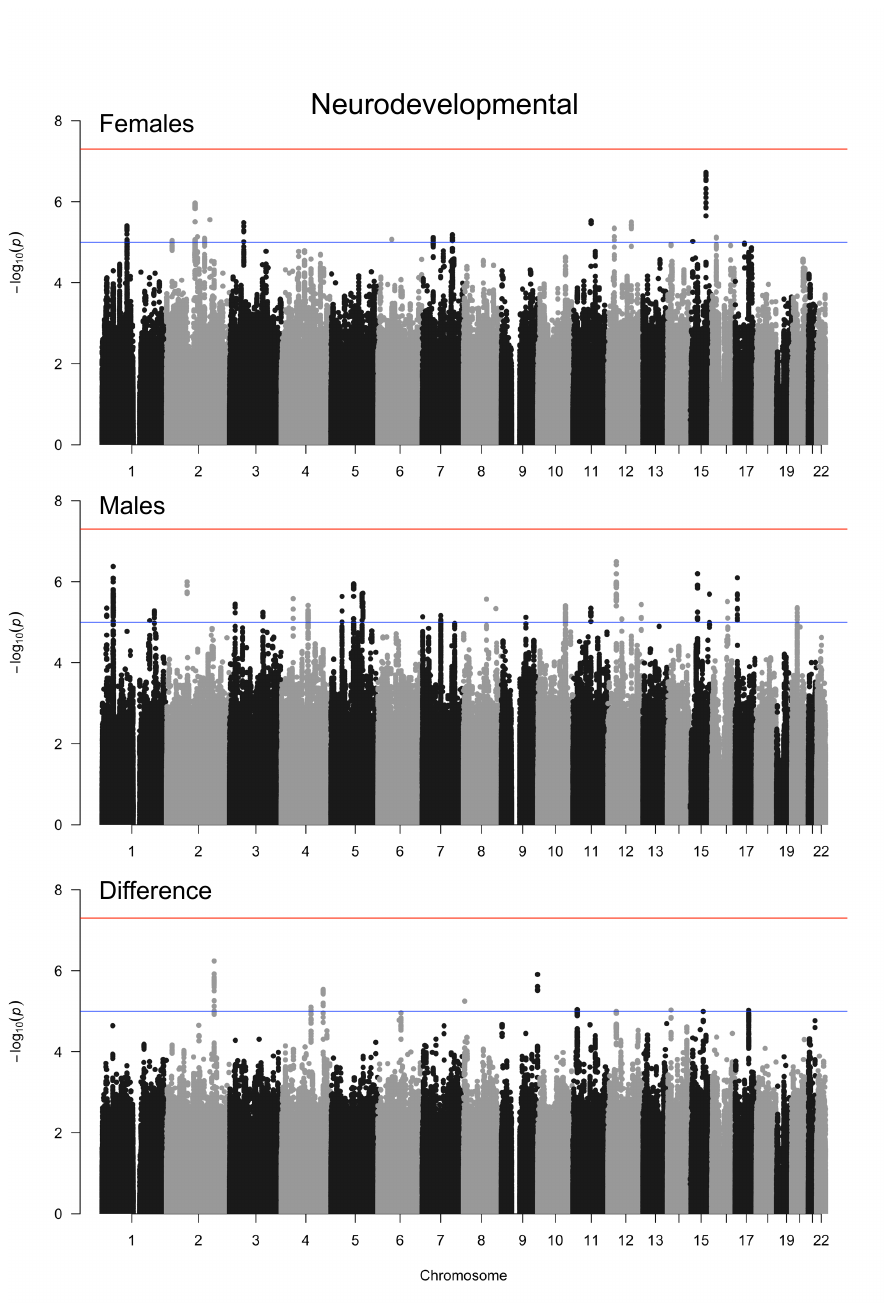

**Figure S5. Manhattan plots of sex differentiation in Neurodevelopmental factor associations across SNPs.** The red line denotes the threshold for genome-wide significance ($5\times{10}^{-8})$ and the blue line denotes the threshold for suggestive genome-wide associations ($5\times{10}^{-6}$). Data were available for 4.53 million SNPs.

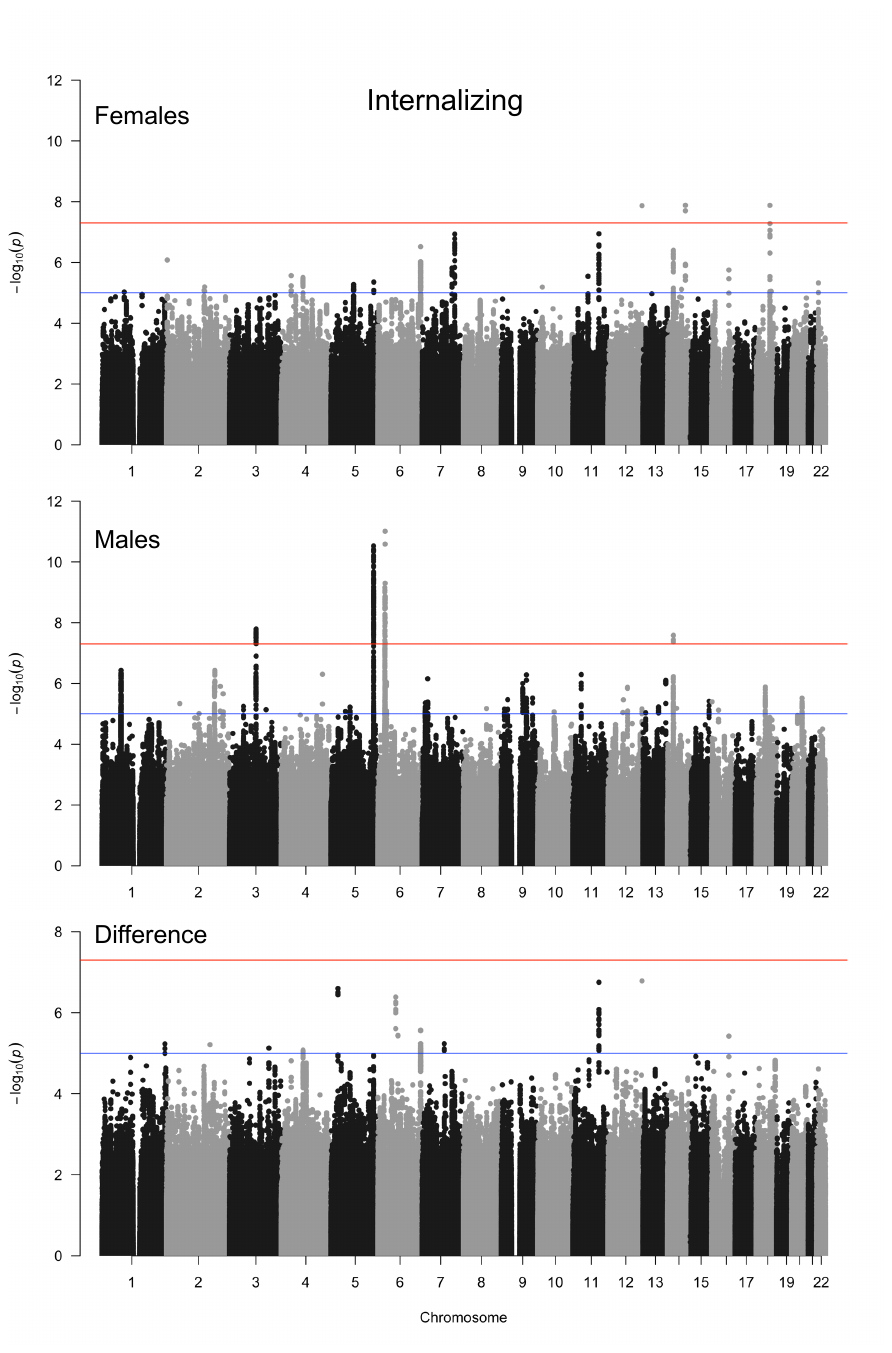

**Figure S6. Manhattan plots of sex differentiation in Internalizing factor associations across SNPs.** The red line denotes the threshold for genome-wide significance ($5\times{10}^{-8})$ and the blue line denotes the threshold for suggestive genome-wide associations ($5\times{10}^{-6}$). Data were available for 4.53 million SNPs.

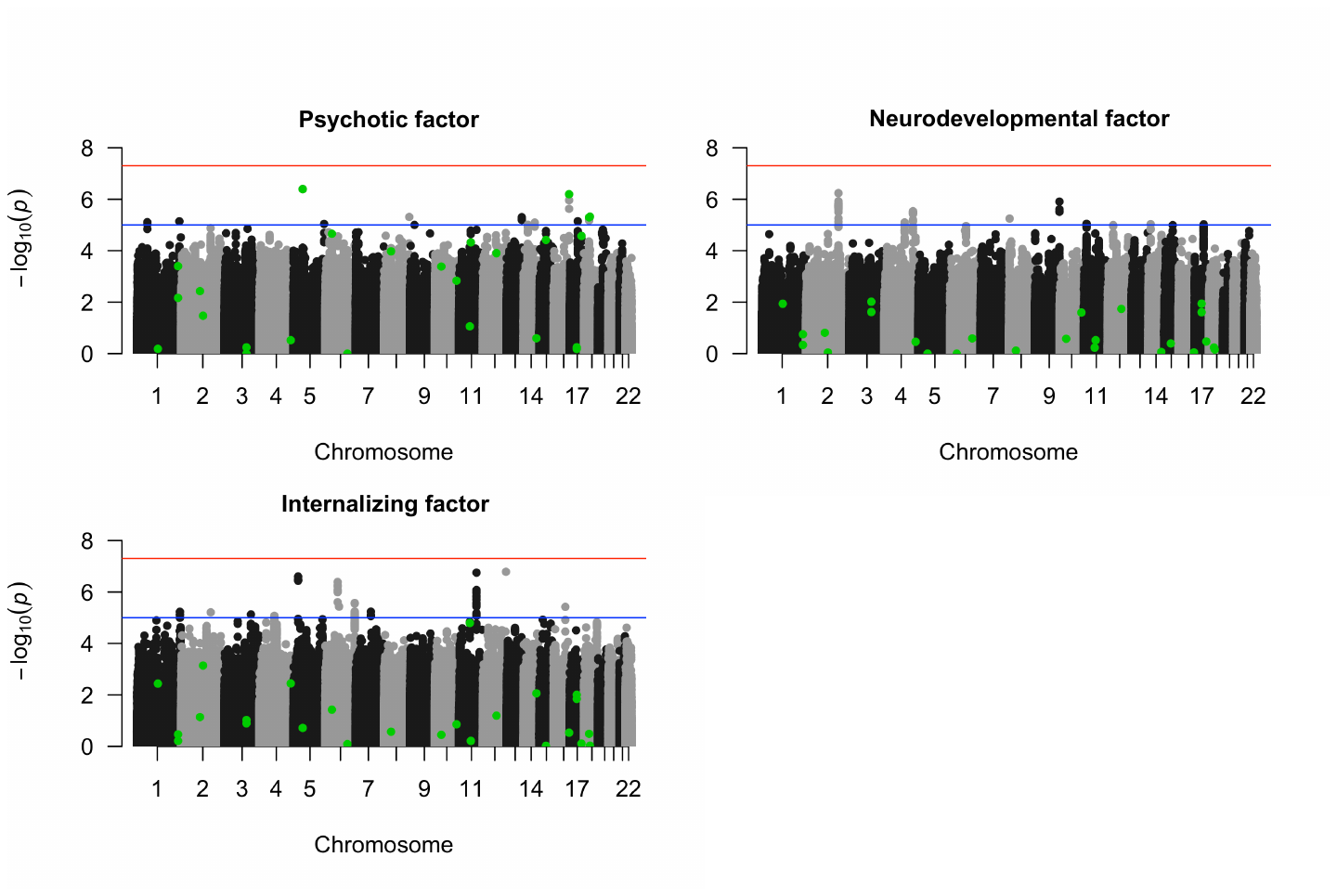

**Figure S7. Sex by SNP interactions, highlighting SNPs that displayed suggestive evidence for sex-differentiated effects on PSY, BIP, MDD, and MTAG-combined PSY, BIP, and MDD in Blokland and colleagues (2022).**

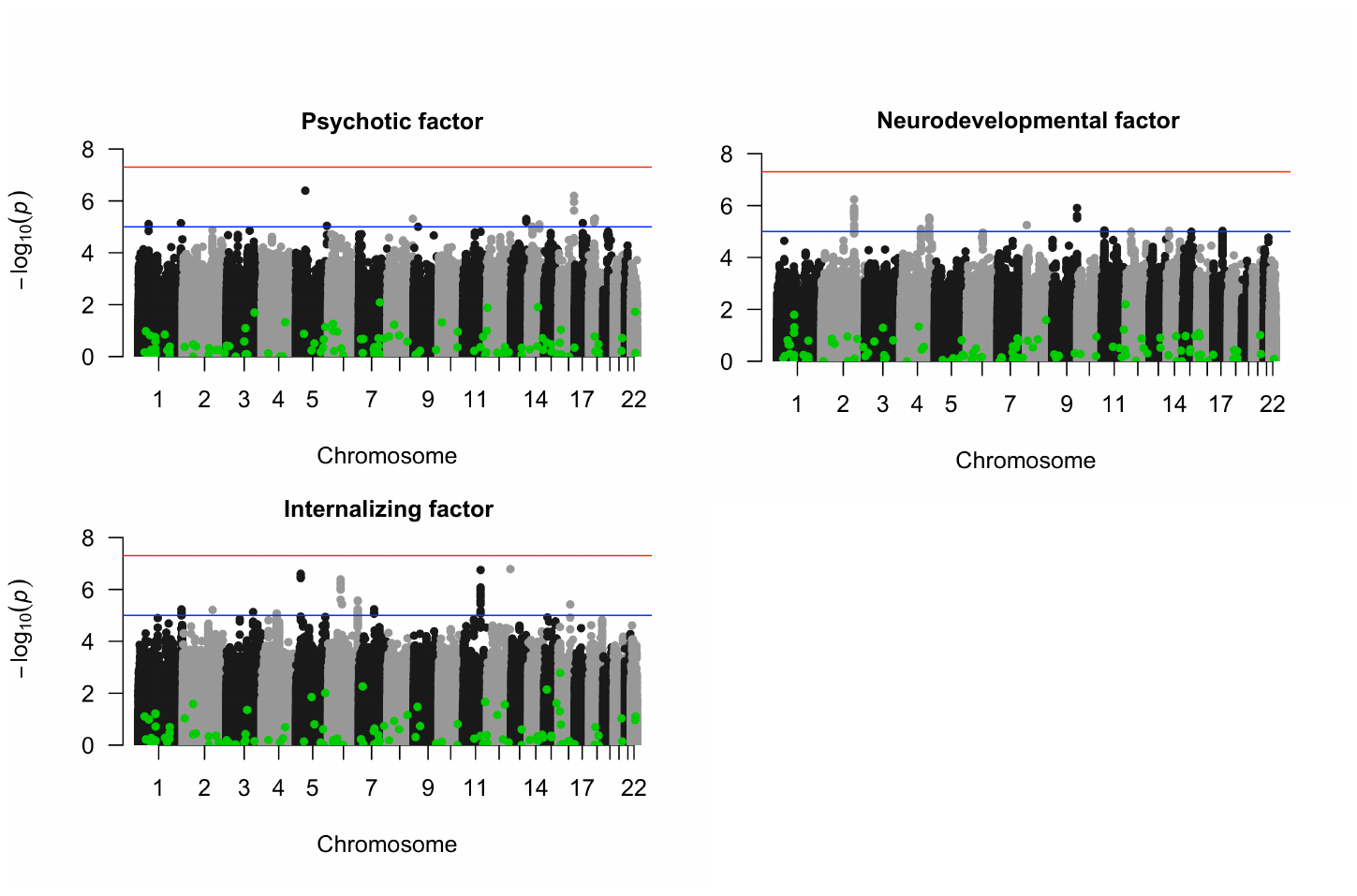
 **Figure S8. Sex by SNP interactions, highlighting SNPs that were genome-wide significant in Grotzinger and colleagues’ (2022) sex-pooled analyses of the multivariate genetic architecture of psychiatric disorders.**

**Sensitivity analyses: Removing the residual between ADHD and ALCH in females**

As the goal of the present study was to identify sex differences in the genetic factor structure of psychiatric disorders, we took precautions to identify and correct for any differences at the level of specific disorders that may not generalize to the factor level. Upon inspection of the genetic covariance and correlation matrices produced by LDSC (**Figures S1 and S2** above), we identified that the genetic association between ADHD and ALCH in females was highly discrepant from that in males. To account for this, we estimated models that included an additional parameter allowing for covariance between the residual error of these two phenotypes. Sensitivity tests comparing the final model estimated with this residual covariance to the final model estimated without indicated little substantive effect of this decision. Model fit was not significantly different (χ^2^(1) = 1.56, *p* = .212), and correlations between factors were only trivially different in the model where this residual was not estimated (change in *r* ≤ .02).

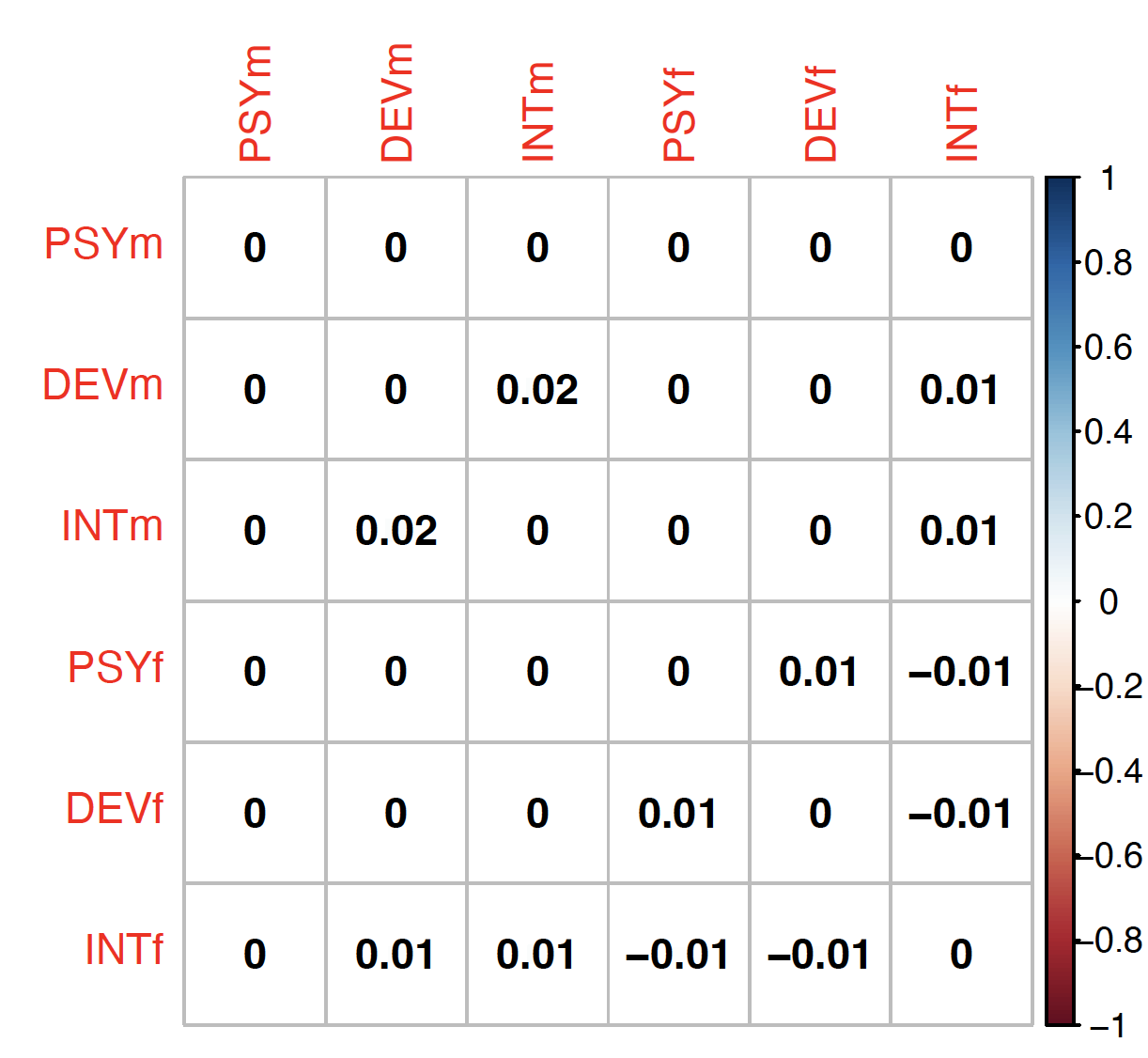

**Figure S9. Change in factor correlations when the residual correlation between ADHD and ALCH in females is not estimated.** Change calculated by subtracting the final model psi matrix from the psi matrix of the model estimated without the correlation.

**Sensitivity analysis: Removing autism in males from the model**

Because univariate LDSC performed on the female sex-stratified autism summary statistics indicated greater intercept inflation than genetic signal, as well as negative heritability (**Table 1**), we did not include this phenotype in analyses as pre-registered. However, we retained the autism phenotype in the male portion of the model, as it can provide additional genetic signal to more precisely estimate the neurodevelopmental phenotype. Sensitivity analyses comparing the final model estimated with autism in males to the final model with autism omitted indicated substantive effects of this change: removing AUT_m_ decreased precision for estimation of the heritability of the common variance underlying the neurodevelopmental factor (SE of factor SD increased from .032 to .188), and removing autism from the model substantially *decreased* correlations between the neurodevelopmental factor in males and all other disorder factors, as shown in **Figures S10** and **S11** below. With autism removed from the model, sex differentiation in associations between the psychotic factor and the neurodevelopmental factor was attenuated but persisted (in males, *r* = .20, in females, *r* = .05, χ^2^(1) = 4.00, *p* = .045).

Because including autism in the model substantially increased the correlation between the neurodevelopmental factor in males and females (from *r* = .86 to *r* ~ 1), we believe that including autism in the model thus makes our estimates of sex differences in the multivariate genetic architecture of psychiatric disorders more conservative in nature, as the neurodevelopmental factors estimated in males and females are paradoxically made more similar in content when modeled with different indicators. We discuss this result further in the discussion section of the main text.

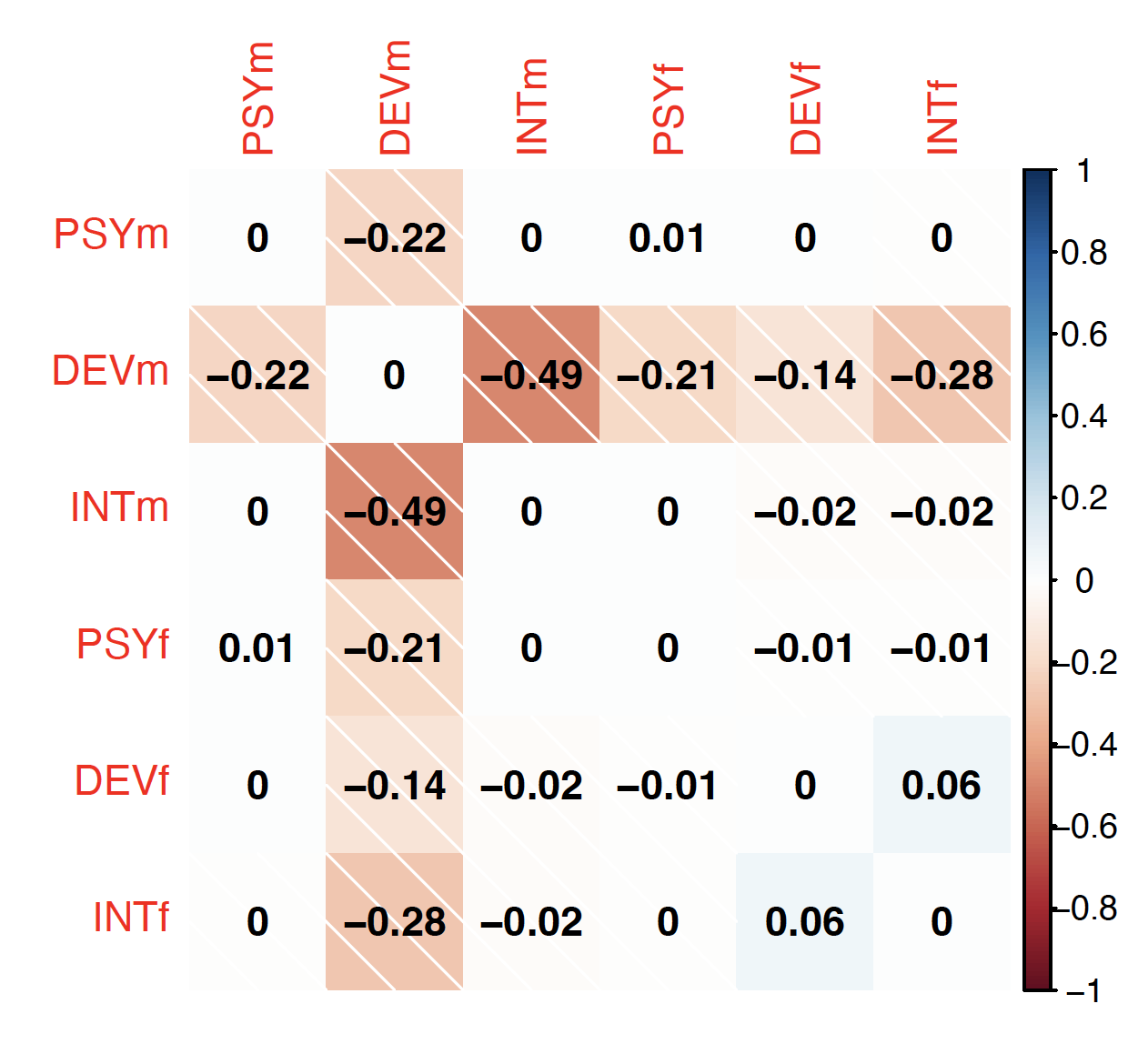

**Figure S10. Change in factor correlations when removing autism in males from the model.** Change calculated by subtracting the final model latent variable covariance matrix from the latent variable covariance matrix of the model estimated without autism in males.

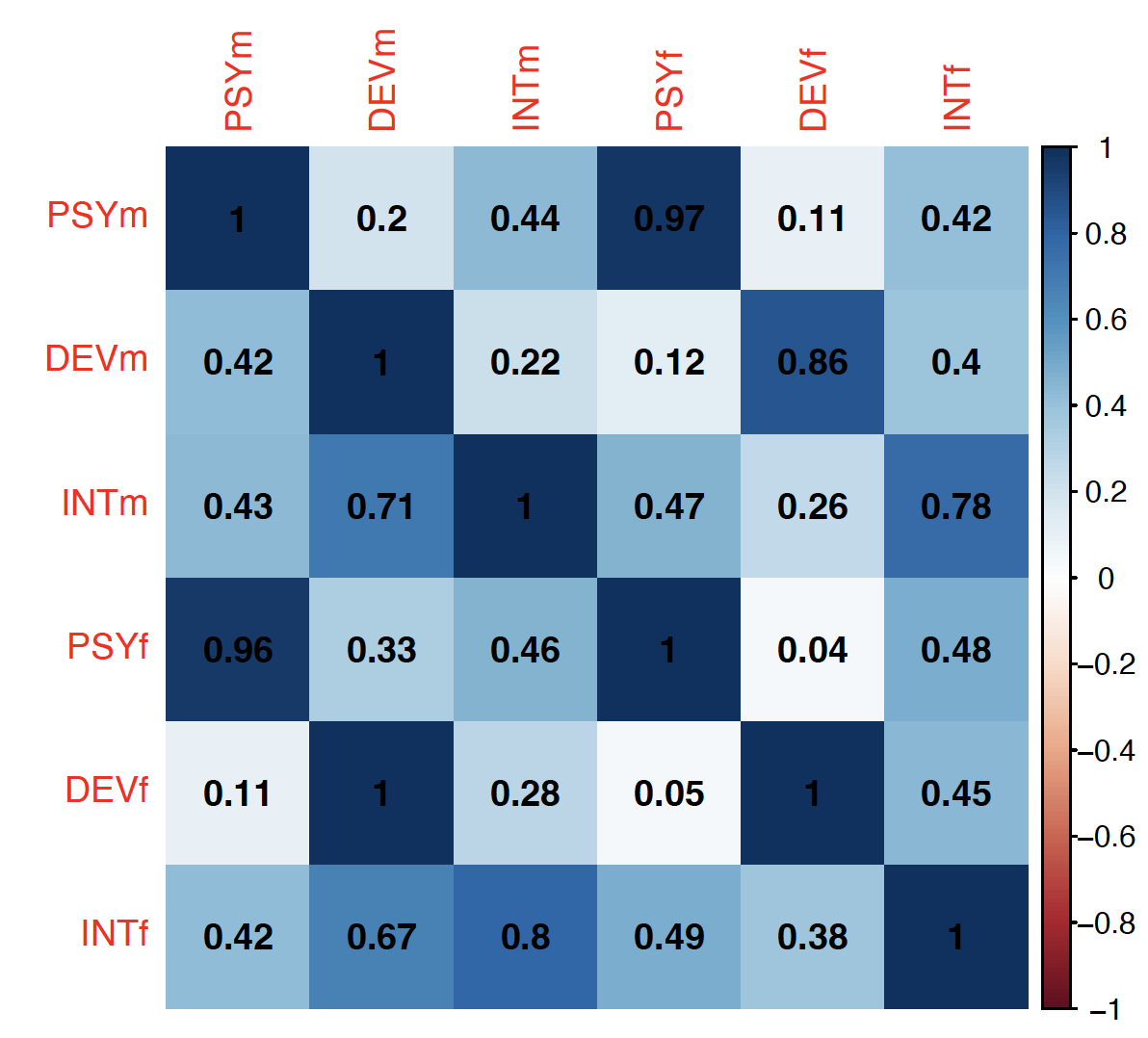

**Figure S11. Factor correlations for the final model including AUT_m_ (below diagonal) and excluding AUT_m_ (above diagonal)**

**Sensitivity analysis: Adding a correlated residual with ALCH in tests of sex-differentiated outcome associations**

As factor loadings on ALCH differed substantially across males and females, we examined whether the four significant sex-differentiated associations between outcomes and the psychotic factor were driven by this difference. To do this, we estimated the sex-differentiated outcome models as described in the main text with one change: an added correlated residual from the outcome variable measured in each sex to ALCH in each sex. For each of the four outcomes (smoking ever, Townsend deprivation index, educational attainment, and insomnia), correlations with the psychotic factor were still significantly different between males and females, as indicated by nested model comparison test (all *p*s < .0002), and each difference remained smaller in magnitude than the localSRMD cutoff of .150 for substantial differences (**Table S5**).

**Group differentiation in genetic associations when some data are pooled across groups**

Genetic associations between two phenotypes can be estimated using LDSC regardless of the sample overlap between those phenotypes. This allows researchers to test group differences even when some GWAS summary data are pooled across groups. For example, instead of comparing the genetic correlation between internalizing disorders and neuroticism in males with the genetic correlation between internalizing disorders and neuroticism in females, a researcher could compare whether internalizing disorders measured in males or internalizing disorders measured in females are more strongly associated with neuroticism measured in a sample of both males and females. Investigating group differences in this way may be practically useful, as GWAS results are often pooled across many subgroups (e.g., participants are not stratified by sex, SES, or age) and so it may be difficult to obtain group-stratified summary statistics for all phenotypes of interest.

We investigated whether group differences that can be identified when data are group-stratified are also identifiable when data are group-pooled, using statistical simulation and then real data. First, we considered a simple hypothetical data-generating model (**Figure S12 panel A**), where a disorder and an outcome were measured separately in 500,000 males and 500,000 females. The outcome was simulated to genetically correlate .80 across sex, similar to phenotypes like waist-hip ratio and heart problems that show a moderate amount of sex-differentiated genetic architecture (c.f. Fig. 2 of Bernabeu et al., 2021). In males, the correlation between the disorder and the outcome was simulated to be .20, and in females, the correlation was .50, representing a true sex difference of *r* = .30 (which would roughly correspond to a localSRMD of .15, our pre-registered cutoff for a meaningfully large sex difference in factor-outcome associations).

We expect genetic associations between a sex-differentiated disorder and a sex-pooled outcome to be a weighted mean of the genetic association between the within-sex disorder-outcome association and the between-sex disorder-outcome association, with weights corresponding to the relative size of the male and female subsamples. We express this in Equation S1 for males:

$$cov\left( {DIS}_{m},{OUT}_{mf} \right)=\frac{w_{outm}*cov\left( {DIS}_{m},{OUT}_{m} \right)+w_{outf}* cov\left( {DIS}_{m},{OUT}_{f} \right)}{w_{outm}+w_{outf}}$$

And in Equation S2 for females:

$$cov\left( {DIS}_{f},{OUT}_{mf} \right)=\frac{w_{outf}*cov\left( {DIS}_{f},{OUT}_{f} \right)+w_{outm}* cov\left( {DIS}_{f},{OUT}_{m} \right)}{w_{outm}+w_{outf}}$$

where phenotypes subscripted *m* are measured in males, *f* in females, and *mf* in a combined sample of males and females, and *w* is the weight for the subscripted variable.

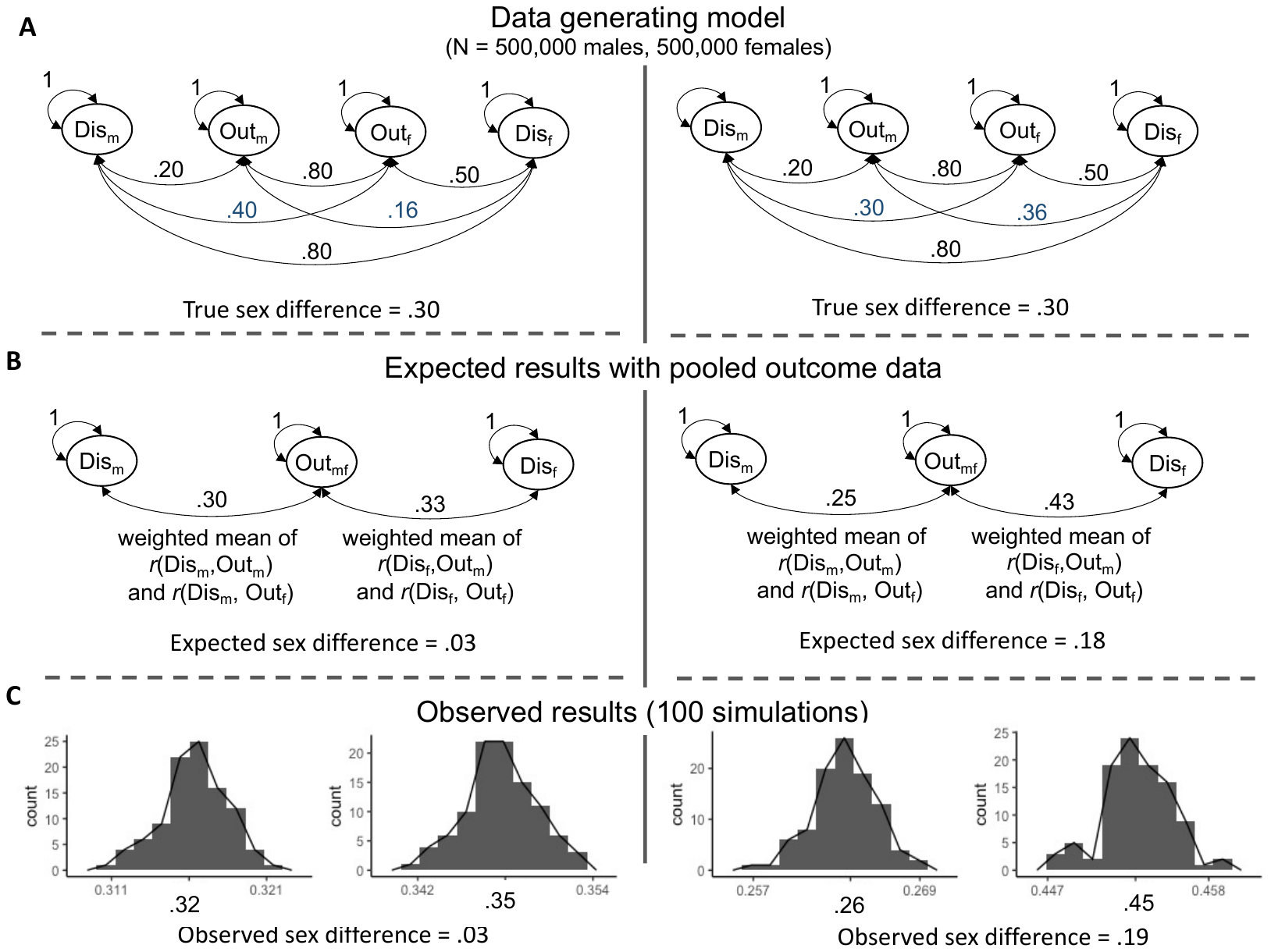

**Figure S12. Sex differences in genetic associations with sex-pooled outcomes: two hypothetical scenarios.** All parameters in the left and right data generating models are the same except for the cross-sex disorder-outcome correlations, which are depicted in blue in the middle row in panel A. Due to these differences, In the left case, true sex differences are not recovered with sex-pooled outcome data (true sex difference = .30, observed sex difference = .03). In the right case, true differences are recovered, albeit with reduction in effect size (true sex difference = .30, observed sex difference = .19).

As we illustrate in **Figure S12 panel B**, whether sex differentiation in genetic associations can be identified with sex-pooled data is contingent on whether cross-sex and within-sex disorder-outcome differences are congruent with one another. In the left half of the figure, the within-sex disorder-outcome correlation is .30 greater in females, but the cross-sex correlation is .24 *smaller* in females. As these differences are averaged together when outcome data are pooled, true sex differences in the data generating model will be obscured, leading to an expected sex difference of *r* = .03 in sex-pooled outcome data. In the right half of the figure, however, the within-sex disorder-outcome correlation is *r* = .30 greater in females, whereas the cross-sex disorder-outcome correlation is only *r* = .06 smaller. As there is greater concordance between these two differences, the true sex differences are better represented in the sex-pooled results: we would expect to find a sex difference of *r* = .18.

To confirm these predictions, we simulated genetic associations according to the data generating model in **Figure 12 panel A** using the simLDSC function found in the development version of GenomicSEM (de la Fuente et al., 2022). To create sex-pooled outcome data, we meta-analyzed the male-only and female-only results into a set of sex-pooled summary statistics using a fixed-effects, inverse-variance weighted model. Across 100 simulations, the results of these simulated data (**Figure S12 panel C**) match our expected results in panel B: In the left model, the true sex differences in the data-generated model are not identified in the sex-pooled model, whereas in the right model, they are, though the expected difference is smaller than the true difference.

We tested whether these differences could be recovered in real data, using the four outcomes from our study that had significant sex-differentiated associations in sex-stratified data: smoking ever, educational attainment, insomnia, and Townsend Deprivation Index. For each of these variables, we meta-analytically combined summary statistics across males and females in the same way we combined simulated summary statistics above. We then tested whether these combined summary statistics showed significantly differentiated associations with the psychotic disorders factor.

For both smoking ever (*p* = 1.59*10^-7^$)$and educational attainment (*p* = 1.19*10^-7^) we were able to identify significantly sex-differentiated associations even when the phenotype was measured in a sex-combined sample (**Table S6**). However, we were unable to recover differentiated associations with insomnia (*p* = .006) and Townsend Deprivation Index (*p* = .452), as within-sex disorder-outcome differences did not extend to cross-sex disorder-outcome differences. As identified in simulations above and **Table S7**, this is because for smoking ever and educational attainment, the sex-pooled association reflects a weighted combination of two very similar sex-stratified associations (equation S1 and S2), which preserves the sex-stratified differences. For example, in sex-stratified data, *r*(Smoking Ever_m_, PSY_m_) = .238 and *r*(Smoking Ever_f_, PSY_f_) = .087, a difference of *r* = .151, and in pooled outcome data, *r*(Smoking Ever_mf_, PSY_m_) = .198 and *r*(Smoking Ever_mf_, PSY_f_) = .085, a similar difference of *r* = .113. However, for insomnia and deprivation, the sex-pooled association reflects a weighted combination of discrepant sex-stratified associations, which obfuscates sex-stratified differences. For example, in sex-stratified data, *r*(Insomnia_m_, PSY_m_) = .112 and *r*(Insomnia_f_, PSY_f_) = -.048, a difference of *r* = .150, but in pooled outcome data, *r*(Insomnia_mf_, PSY_m_) = .039 and *r*(Insomnia_mf_, PSY_f_) = .023, a difference of only *r* = .016.

Overall, these results indicate that some, but not all, group-differentiated genetic associations can be identified when one phenotype in the genetic association is not measured with group stratification, although the recovered difference is smaller than the true difference. In statistical language, there is an inflated type-II error rate when conducting these tests, and thus null group differences should be interpreted conservatively (further simulation and testing is needed to understand how type-I error rate is affected by this method).

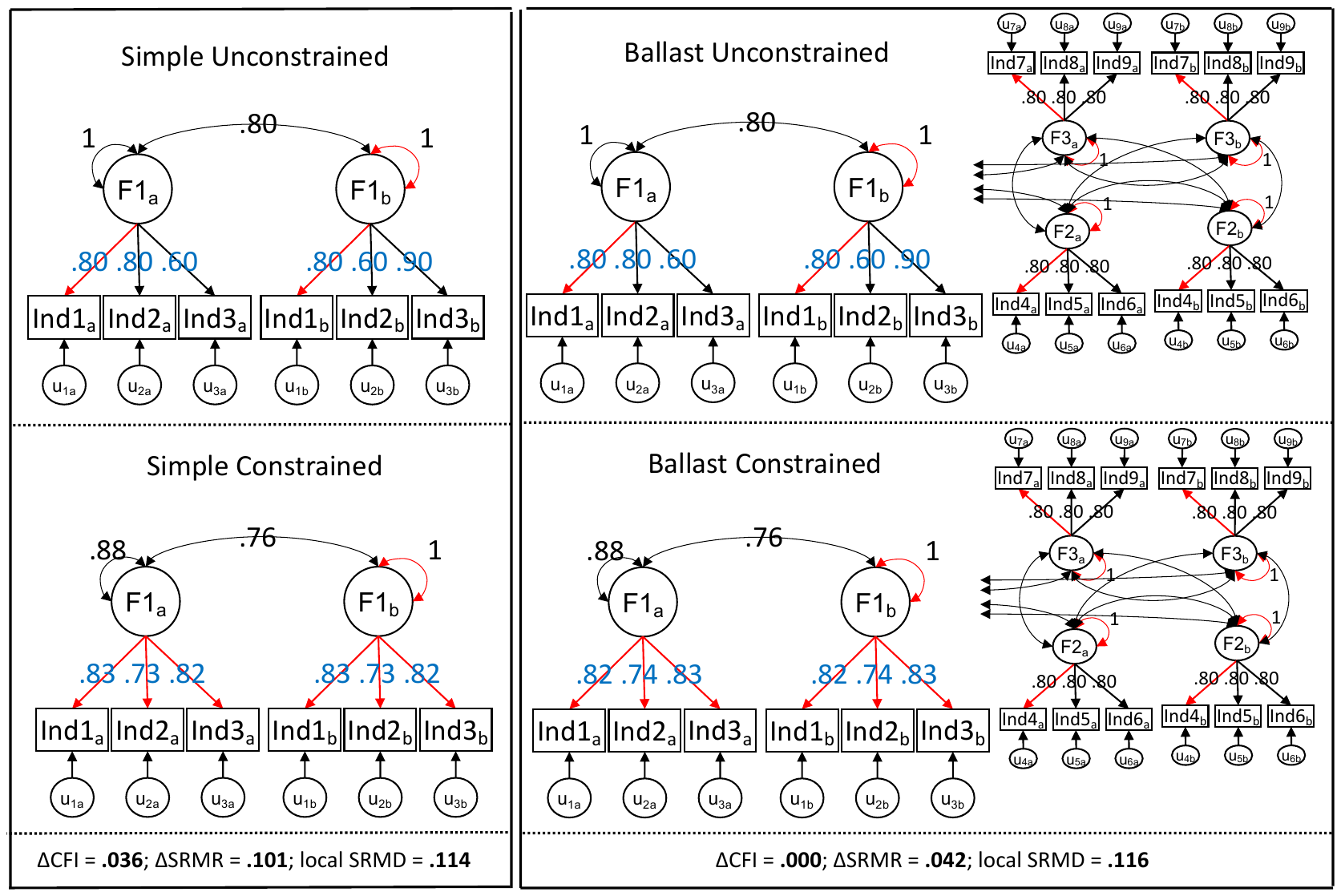

**Figure S13. A demonstration of how model ballast affects CFI and SRMR but not localSRMD.**

This figure depicts two sets of simulated multigroup models: the left side depicts simple models, where one latent factor (F1) is measured in two groups (a and b) using three indicators (Ind1-3). The right side depicts more complex models, where, in addition to F1, two additional “ballast” factors are added (F2 and F3; left-facing covariance paths from F2 and F3 in the ballast models represent covariances with F1). Comparison between these two sets of models illustrates the ability of different fit indices to detect localized model misfit. Specifically, in both the simple and ballast models, latent factor F1 has different patterns of factor loadings in groups a and b (blue numbers). In both sets of models, adding constraints (red parameters) that force factor loadings to be equal across groups a and b introduces misfit. In the simple models, this misfit can be detected with each of CFI, SRMR, and localSRMD (bottom of figure). In the ballast models, this misfit is only detected using localSRMD. The reason for this discrepancy is that additional factors estimated in the ballast model (F2 and F3) remain unchanged across the unconstrained and constrained models, which reduces the sensitivity of global fit indices like CFI and SRMR to detecting misfit, but these differences do not affect the sensitivity of localSRMD.

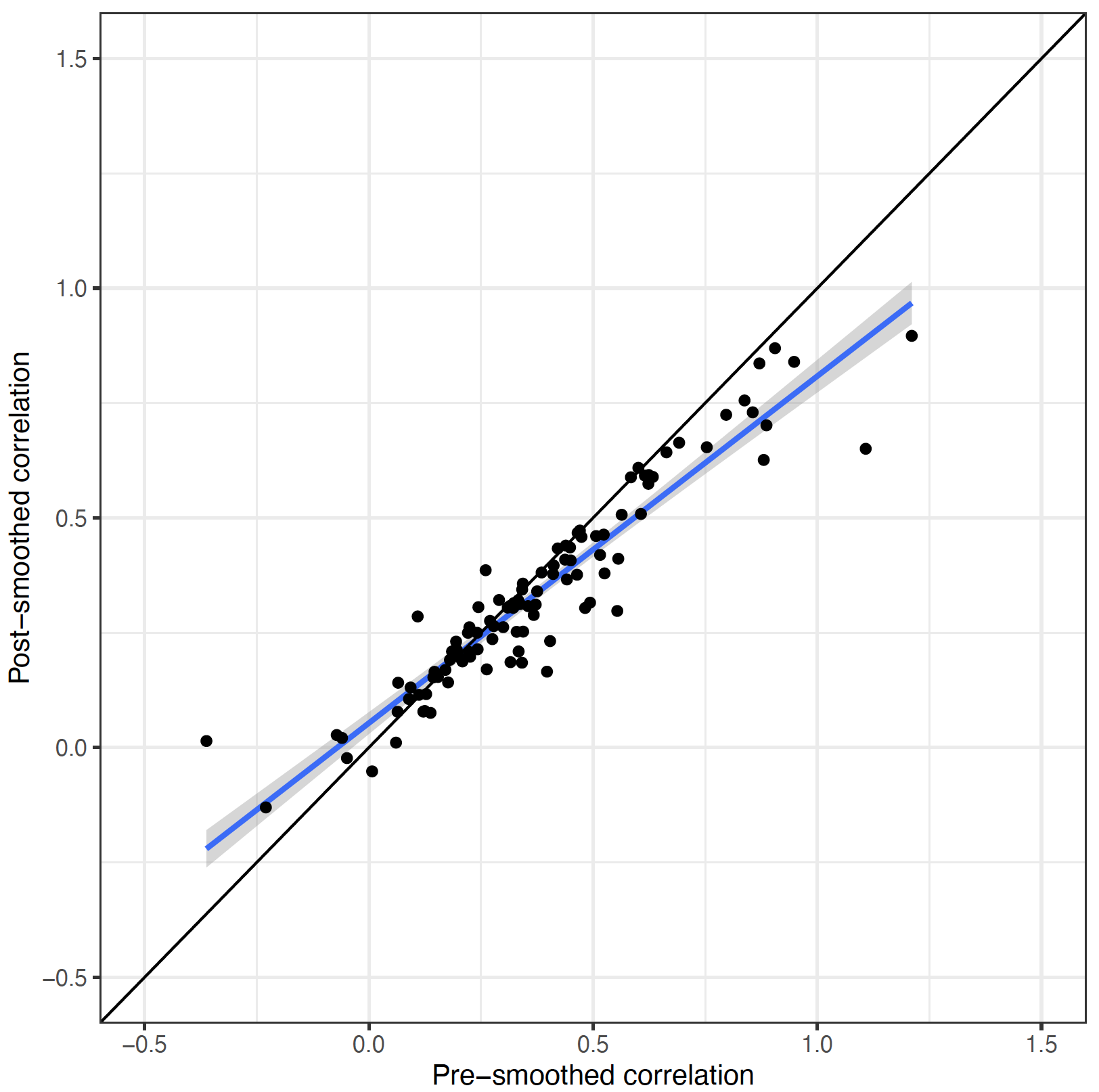

**Figure S14. Comparison of pre- and post-smoothed genetic covariance matrices, in terms of correlations.**

**Supplemental References not cited in main manuscript**

Chang, C. C., Chow, C. C., Tellier, L. C., Vattikuti, S., Purcell, S. M., & Lee, J. J. (2015). Second-generation PLINK: rising to the challenge of larger and richer datasets. *Gigascience*, *4*(1), s13742-015.

de la Fuente, J., Grotzinger, A. D., Marioni, R. E., Nivard, M. G., & Tucker-Drob, E. M. (2021). Multivariate Modeling of Direct and Proxy GWAS Indicates Substantial Common Variant Heritability of Alzheimer's Disease. *medRxiv*.

Howard, D. M., Adams, M. J., Clarke, T. K., Hafferty, J. D., Gibson, J., Shirali, M., ... & McIntosh, A. M. (2019). Genome-wide meta-analysis of depression identifies 102 independent variants and highlights the importance of the prefrontal brain regions. *Nature Neuroscience*, *22*(3), 343-352.

Hyde, C. L., Nagle, M. W., Tian, C., Chen, X., Paciga, S. A., Wendland, J. R., ... & Winslow, A. R. (2016). Identification of 15 genetic loci associated with risk of major depression in individuals of European descent. *Nature genetics*, *48*(9), 1031-1036.

Psychiatric GWAS Consortium Bipolar Disorder Working Group (2011). Large-scale genome-wide association analysis of bipolar disorder identifies a new susceptibility locus near ODZ4. *Nature Genetics*, *43*, 977–983.
